# Online sexual, reproductive, and transgender healthcare for LGBTQI+ youth: A scoping review

**DOI:** 10.1101/2024.08.06.24311547

**Authors:** Julie McLeod, Claudia S. Estcourt, Paul Flowers, Jo Gibbs, Jennifer MacDonald

**Author notes:** These authors contributed equally to this manuscript (Claudia S. Estcourt, Paul Flowers).

## Abstract

**Background:** Lesbian, gay, bisexual, trans, queer, questioning, intersex, and other sexual and gender minority (LGBTQI+) youth have poor sexual and reproductive health outcomes and low uptake of sexual and reproductive healthcare (SRHC). Online SRHC and transgender healthcare could overcome known barriers to in-person SRHC, such as confidentiality concerns. Therefore, we aimed to describe existing literature on online SRHC and transgender healthcare for LGBTQI+ youth, synthesise study findings, and make recommendations for future research.

**Methods:** We conducted a scoping review following the Joanna-Briggs Institute methodology. Eligibility were online SRHC and transgender healthcare for LGBTQI+ youth (aged 10-35 years) in high-income countries. Search strings were framed around the eligibility criteria and 265 search terms were selected to identify published literature from nine databases. Searches were exported to Rayyan and studies screened by two reviewers. Data from included studies were extracted to Excel and analyzed descriptively.

**Results:** Of 91 included papers, 41 were quantitative, 26 were qualitative, and 24 were mixed methods. Seventy-one papers focused on sexual health (HIV/sexually transmitted infection (STI) prevention 52/71; HIV management 9/71; sexual health *per se* 9/71; and HIV stigma reduction 2/71); 3/91 on reproductive health (pregnancy prevention 2/3); 2/91 sexual and reproductive health; and 16/91 on transgender health (gender identity/transition *per se* 8/16; gender affirming care 8/16). Papers explored the provision of or engagement with education/information (72/91); non-clinical support (56/91, e.g., reminders for HIV/STI testing); and clinical care (18/91) for sexual health (10/18, e.g., home HIV/STI self-sampling kits 6/10) or transgender health (8/18, i.e., eConsultation with a healthcare provider 8/8). Studies targeted young men who have sex with men (62/91) for sexual health; trans and gender diverse youth (26/91) for transgender healthcare (16/26) and sexual health (14/26); LGBTQI+ youth (6/91); and young sexual minority women (4/91) for reproductive health (3/4) and sexual and reproductive health (1/4).

**Conclusions:** There is a large and varied literature base for online SRHC and transgender healthcare for LGBTQI+ youth. However, most research focused on sexual healthcare, particularly HIV/STI prevention, for men who have sex with men. Very little explored reproduction or sexual health other than HIV prevention. Young sexual minority women and trans and gender diverse youth are notably under-researched for online SRHC. Research is needed to understand how to enhance the potential of online healthcare for LGBTQI+ youth.

## Introduction

Lesbian, gay, bisexual, trans, queer, questioning, intersex, and other sexual orientation and gender diverse (LGBTQI+) youth (e.g., aged 10-35 years) face a disproportionate burden of poor sexual and reproductive health, including high rates of sexually transmitted infections (STIs) and blood borne viruses (BBVs), sexual violence and abuse, low sexual wellbeing, and unplanned pregnancy at a young age (1–10) (see S1 Appendix for definitions of key terms used throughout this paper). In particular, young gay, bisexual and other men who have sex with men (GBMSM) and trans women who have sex with men are at higher risk for STIs and BBVs (1–5) and young bisexual women and trans youth are at higher risk for unplanned pregnancy and sexual violence and abuse (1–3,5,7–9). Additionally, LGBTQI+ youth have low uptake of sexual and reproductive healthcare (SRHC), such as testing for STIs/BBVs, getting vaccinated for human papilloma virus, and uptake of pre-exposure prophylaxis (PrEP) (11–16). LGBTQI+ youth also face considerable barriers to engaging with care (1,2), including confidentiality concerns, lack of perceived risk or necessity, discrimination and stigma, healthcare providers’ lack of knowledge and training about their needs, and heteronormative assumptions of their gender and sexuality (17–21). When considering these inequalities, it is important to be cognizant of the intersectionality of gender and sexual orientation with other socio-economic demographics/characteristics associated with increased risk of poor outcomes, such as race/ethnicity and socio-economic status (22–24). In the United Kingdom (UK) and United States of America (USA), young Black GBMSM bear the largest burden of HIV incidence (6,25,26). Further, stigma experienced by young LGBTQI+ people of color in the USA and Canada can impact identity disclosure to healthcare provider and decisions about uptake of PrEP (27). The delivery of SRHC online (i.e., via websites, mobile apps, web apps) has the potential to increase uptake of sexual and reproductive health and improve sexual and reproductive health outcomes among LGBTQI+ youth by overcoming the barriers to in-person services (28–33). However, research into online SRHC innovations often focuses on general populations rather than specifically LGBTQI+ youth (e.g., 34–38), risking online services not meeting the needs of LGBTQI+ youth.

In addition, for trans and gender diverse youth, a critical issue for sexual and reproductive health is transgender health. Based on the premise that gender and sex are distinct (39) and some individuals can experience emotional distress due to incongruence between their gender and sex (40), known as gender dysphoria, transgender health refers to the ability for trans and gender diverse individuals to live in the gender that feels most authentic and comfortable. Gender dysphoria and related anxiety and depression can negatively impact trans and gender diverse people’s uptake of SRHC (41,42). Equally, receiving gender affirming care (medical care such as hormones and/or surgery to affirm one’s gender and align their body and gender identity) is also associated with increased use of SRHC among trans and gender diverse youth, such as STI testing and awareness of PrEP (43). However, trans and gender diverse youth also face barriers to gender affirming care, in particular, long wait times (typically years for a consultation) (44–46). Furthermore, transgender healthcare (broader than gender affirming care, including information, support, and clinical care regrading gender identity, expression, and transition (47)) is also intrinsically interlinked with reproductive healthcare, as a key aspect of transgender healthcare can be fertility preservation or assistance (48,49). Thus, the delivery of integrated SRHC and transgender healthcare has the potential to increase uptake of SRHC and improve sexual and reproductive health outcomes among transgender youth. Recent research from Australia found that co-located sexual and reproductive health and gender clinics with both sexual and reproductive health and endocrinology physicians facilitated access to gender affirming care, STI screening, contraception, and cervical screening for trans populations (50). Moreover, transgender women in the USA living with HIV have expressed that integrated HIV care and gender affirming care would be more accessible than current non-integrated services (51). However, SRHC and transgender healthcare are typically delivered and studied separately, and existing research has largely focused on adult populations. The extent of literature into transgender healthcare, delivered both individually and with SRHC, for LGBTQI+ youth is unclear.

Given that online healthcare and the integration of SRHC and transgender healthcare have the potential to overcome key barriers for LGBTQI+, it is critical to understand how online SRHC and transgender health can best be designed and delivered for LGBTQI+ youth. However, online SRHC is broad, covering a wide range of ‘areas’ of sexual and reproductive health (e.g., infection, wellbeing, violence/abuse, fertility), healthcare types (e.g., information, support, and clinical care for prevention, testing, treatment, management, assistance), and online platforms (e.g., websites, mobile apps, web apps). Therefore, the extent and nature of research regarding online SRHC and transgender healthcare for LGBTQI+ youth is unclear. A review is needed to map existing literature and identify where there are gaps in research (52,53) regarding online SRHC and transgender healthcare for LGBTQI+ youth in order to understand where future intervention and service efforts should focus to improve the uptake of online SRHC and sexual and reproductive health outcomes among LGBTQI+ youth. Thus, the objective of this current study was to identify and describe existing literature on online SRHC and transgender healthcare for LGBTQI+ youth, synthesize study findings, and make recommendations for future research. Three research questions (RQs) were addressed:

RQ1: What types of online sexual and reproductive healthcare and transgender healthcare have received attention for LGBTQI+ youth and where are there gaps?

RQ2: Who are the target LGBTQI+ youth populations of online sexual and reproductive healthcare and transgender healthcare research, which additional intersectional characteristics have been considered, and where are there gaps?

RQ3: How, if at all, have theories, models, and frameworks been used in research into online sexual and reproductive healthcare and transgender healthcare for LGBTQI+ youth?

## Methods

### Design

A systematic scoping review in accordance with the Joanna-Briggs Institute (JBI) methodology for scoping reviews (52–54).

### Protocol registration

The protocol was published in medRxiv (55). See S2 Appendix for protocol deviations.

### Eligibility criteria

Using the Participant, Concept, Context (PCC) framework (53), we included research regarding LGBTQI+ youth aged 10-35 years (Participants); online SRHC and transgender healthcare (Concept); and studies from high-income and developed economy countries (56,57) (Context) published in the past five years (2018–2023). See the protocol and S3 Appendix for detailed inclusion and exclusion criteria and their rationale.

### Types of sources

Only published literature was included. Qualitative, quantitative, and mixed methods studies that were classified as original research with primary data collection were included. Studies using theory-based implementation and behavioural science for intervention development and evaluation were also considered for inclusion. Pilot and feasibility studies were included. Reviews, conference abstracts, posters, registered reports, blogs, guidelines, text and opinion papers, letters, editorials, commentaries, protocols, preprints, and doctoral and master’s theses were excluded. Studies published in any language other than English were excluded, due to lack of resources to support translation.

### Search strategy

The PCC framework was used to structure the search, using only Participants and Concept. A preliminary search of MEDLINE, the Cochrane Database of Systematic Reviews, JBI Evidence Synthesis, and BMJ Open was conducted (31.01.2023) to identify articles on the topic. An analysis of the text words contained in the title and abstract of 10 retrieved papers (31,58–61) was conducted to develop a full search strategy of four search strings (Sexual orientation/gender minority; Age; Online; and Type of health care) and 256 search terms (see S4 Appendix). A second search using all search terms was undertaken across nine databases (22.05.2023): APA PsycInfo (ProQuest); APA PsycArticles (ProQuest); CINAHL Complete (EBSCO); MEDLINE (EBSCO); ERIC (EBSCO); British Education Index (EBSCO); Education Database (ProQuest); Computer Science Database (ProQuest); and Web of Science.

### Study selection

All identified citations were exported to excel files and uploaded to Rayyan (62) and duplicates removed. The titles and abstracts of deduplicated studies were screened (100% by JMcL and 3% (n=178) by RO, see Acknowledgements) for assessment against the eligibility criteria. To prioritize the most relevant studies, the titles and abstracts screened by RO were ordered from most to least relevant, using the ‘Compute Ratings’ function within Rayyan which uses artificial intelligence to calculate the probability of inclusion based on decision patterns (62). The full text of studies categorized as included and maybe were then assessed in detail against the eligibility criteria (100% by JMcL and 10% (n=20) by RO). Of the 178 articles screened by both reviewers, there was 85% consistency. The conflicts were resolved through discussion and referral to the protocol.

### Data extraction and analysis

Data from included papers were extracted to excel by JMcL using a data extraction tool adapted (by JMcL) from the JBI Manual for Evidence charting table for data extraction synthesis (53). The tool was adapted and centered around the PCC framework for extracting relevant data for study details, RQ1, RQ2 and RQ3. The tool was piloted and refined using a relevant paper identified from the preliminary search (63). See S5 Appendix for the final data extraction tool and a narrative account of data extraction and analysis. No authors were contacted to request missing or additional data. Following extraction, analysis involved calculating frequency counts and percentages for study details and relevant data regarding each of the research questions. Analyses were then charted on tables and summarized narratively to provide an overview of the included papers for each of the research questions. Table 1 details data extraction for each of the relevant variables presented in the results.

**Table 1.** Details of data extraction for variables presented in the results.

| Research question | Variable | Data extraction/categorization |
| --- | --- | --- |
| RQ1 | Area of health | Papers were deductively categorized as belonging to sexual, reproductive, or transgender health, based on their health outcomes. |
|  | Health outcomes | Papers were inductively categorized as focusing on one or more of HIV prevention; STI prevention; HIV and STI prevention; HIV management; HIV stigma reduction; Pregnancy prevention; Sexual health (per se); Reproductive health (per se); Gender identity/expression; and Gender affirmation/transition, identified from the title, aim, and/or methods. |
|  | Healthcare types | Papers were deductively categorized as focusing on one or more of Education/Information, Non-clinical Support, and Clinical Care, identified from the title, aim, and/or methods (intervention/service description). *Education/Information* refers to services and interventions that impart information only typically to achieve an increase in knowledge (e.g., information about STIs/HIV; contraception; or gender expression and transition). *Non-clinical Support* refers to services and interventions that provide non-clinical emotional or practical support, beyond information, typically to achieve a desired outcome (e.g., peer communication for HIV stigma reduction; reminders for increased PrEP adherence; or skill building for increased skills for partner notification). *Clinical Care* refers to services |
|  |  | and interventions for medical care, specific to testing, diagnosing, treating, and managing sexual and reproductive health issues or gender affirming care (e.g., STI/HIV testing; uptake or maintenance of PrEP; consultations for gender affirming hormones). |
|  | Online platforms | Papers were inductively categorized as focussing on one or more type of online platform, using verbatim terms extracted from the title, abstract, aim, and/or methods (service/intervention description). |
| RQ2 | Target LGBTQI+ youth population | Papers were inductively categorized as targeting young (aged 10-35 years or youth, young LGBTQI+/adult, teen/adolescent) sexual and gender minority and diverse populations, extracted from the title, abstract, aim, and/or eligibility/inclusion criteria. |
|  | PROGRESS+ | Papers were deductively categorized as considering Place of Residence, Race/Ethnicity, Occupation, Gender/Sex, Religion, Education, Socio-Economic Status, Social Networks (PROGRESS), Features of relationships, Time-dependent relationships, Sexual identity/orientation, Age, Disability, (+) and Living with HIV in recruitment. Data were extracted from inclusion/eligibility criteria and recruitment methods. |
| RQ3 | Theory | Papers were inductively categorized as either having reported or not having reported using a theory, identified from the introduction and/or methods and which theory was used, extracted from the introduction and/or methods. |
|  | How theory was used | Papers were deductively categorized as having used theory in a manner that is.... Identified from the introduction and/or methods. |

### Quality appraisal

In line with JBI scoping review methodology (53), a quality appraisal was not conducted. This review mapped the literature and did not analyze nor draw conclusions from the outcomes of studies.

## Results

### Study selection

Of an initial 5,200 hits, 3,432 remained after deduplication. The titles and abstracts of 3,432 papers were screened and categorized as ‘Excluded’ (n=3,194), ‘Maybe’ (n=138) and ‘Potentially Included’ (n=100). Full-text screening of the ‘Maybe’ and ‘Included’ papers identified 91 papers for inclusion (29–32,43,58–61,64–145).

**Figure 1.**
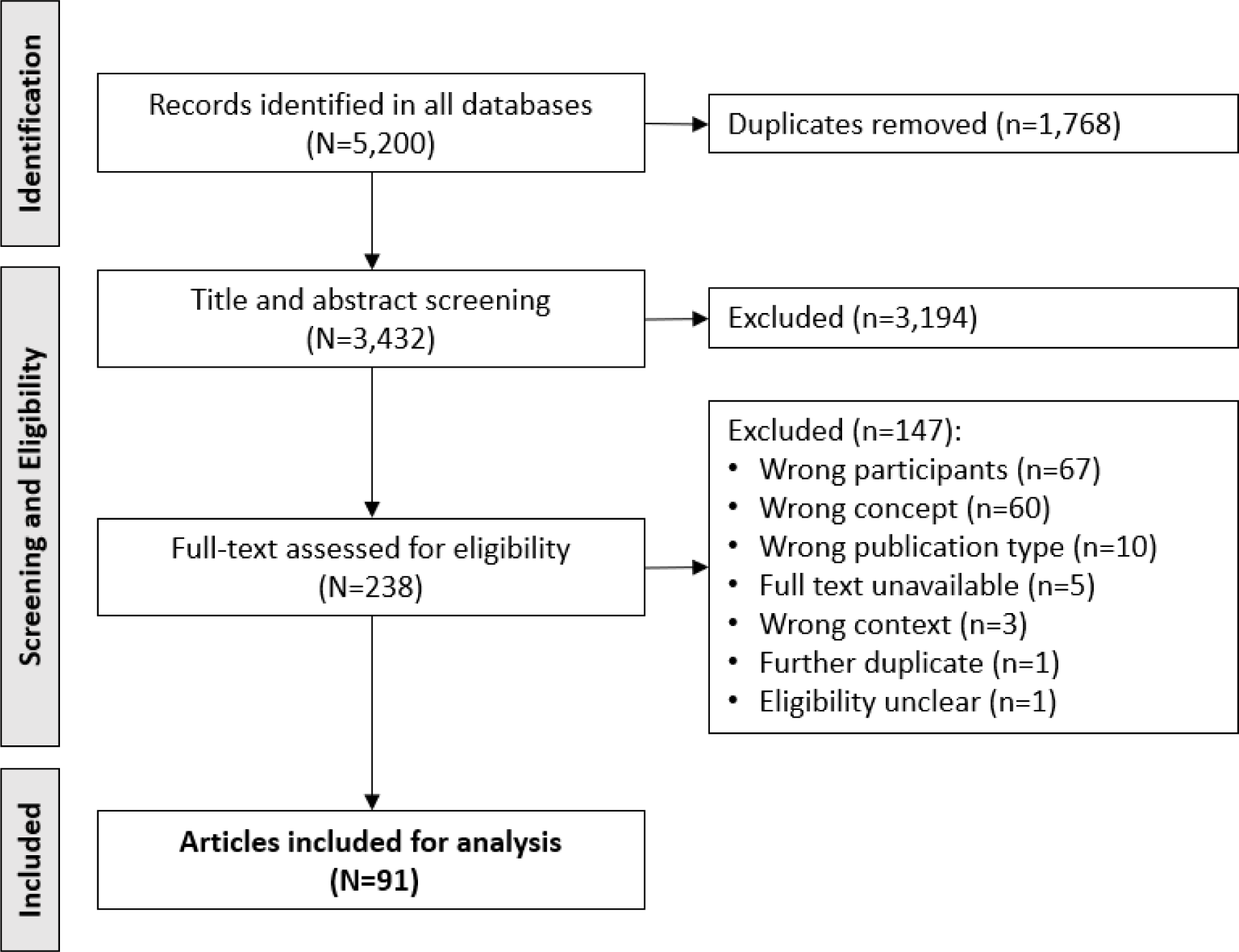
PRISMA-ScR flow chart of study inclusion and exclusion process.

### Study characteristics

Table 2 summarizes the study characteristics of the 91 included papers with citations (for the full dataset, see https://osf.io/ktwxn). Regarding date, the majority of papers were published between 2020 to 2022 (63/91, 69%). Only three papers were published in 2023 (up to May). The vast majority of studies were conducted in the USA (82/91), four in Canada, one in Canada and the USA, two in Australia, one in the Republic of Ireland, and one in the UK (England). Concerning study design and research methods, 41/91 were quantitative, of which 23/41 were randomized control trials (RCTs) and 18/41 were non-RCTs or descriptive - most employed surveys with closed questions (37/41) as their method of data collection; 26/91 papers were qualitative, most of which employed interviews (13/26) or focus groups (8/26); and 24/91 papers reported using both qualitative and quantitative methods, most of which were non-RCTs or descriptive (22/24), largely pairing surveys (21/24) with interviews (11/24) or focus groups (3/24), and/or examining engagement with an intervention, such as number of clicks or length of time spent on it (8/24). Regarding the type of healthcare, the papers either reported on interventions developed for the study (e.g., custom mobile app, novel web app) (62/91) or existing services (e.g., dating apps, government websites) (29/91). Further, most of the papers reported on services or interventions that were real, not hypothetical, meaning the participants could interact with the online platform (84/91), such as ‘the internet’ including websites and social media (e.g., or a mobile app for HIV prevention information and support). A minority were hypothetical (7/91) and sought perspectives on potential services or interventions, such as a mobile app for mentorship regarding HIV prevention. There were 60/91 real interventions, 24/91 real services, 5/91 hypothetical services, and 2/91 hypothetical interventions. Finally, concerning participants, LGBTQI+ youth were participants in most of the papers (82/91); a minority of papers targeted and recruited parents or caregivers (6/91), healthcare providers (4/91), community-based organization staff (1/91) and mentors (1/91) of LGBTQI+ youth. Two papers did not have participants but collected data from existing services for LGBTQI+ youth.

**Table 2.**
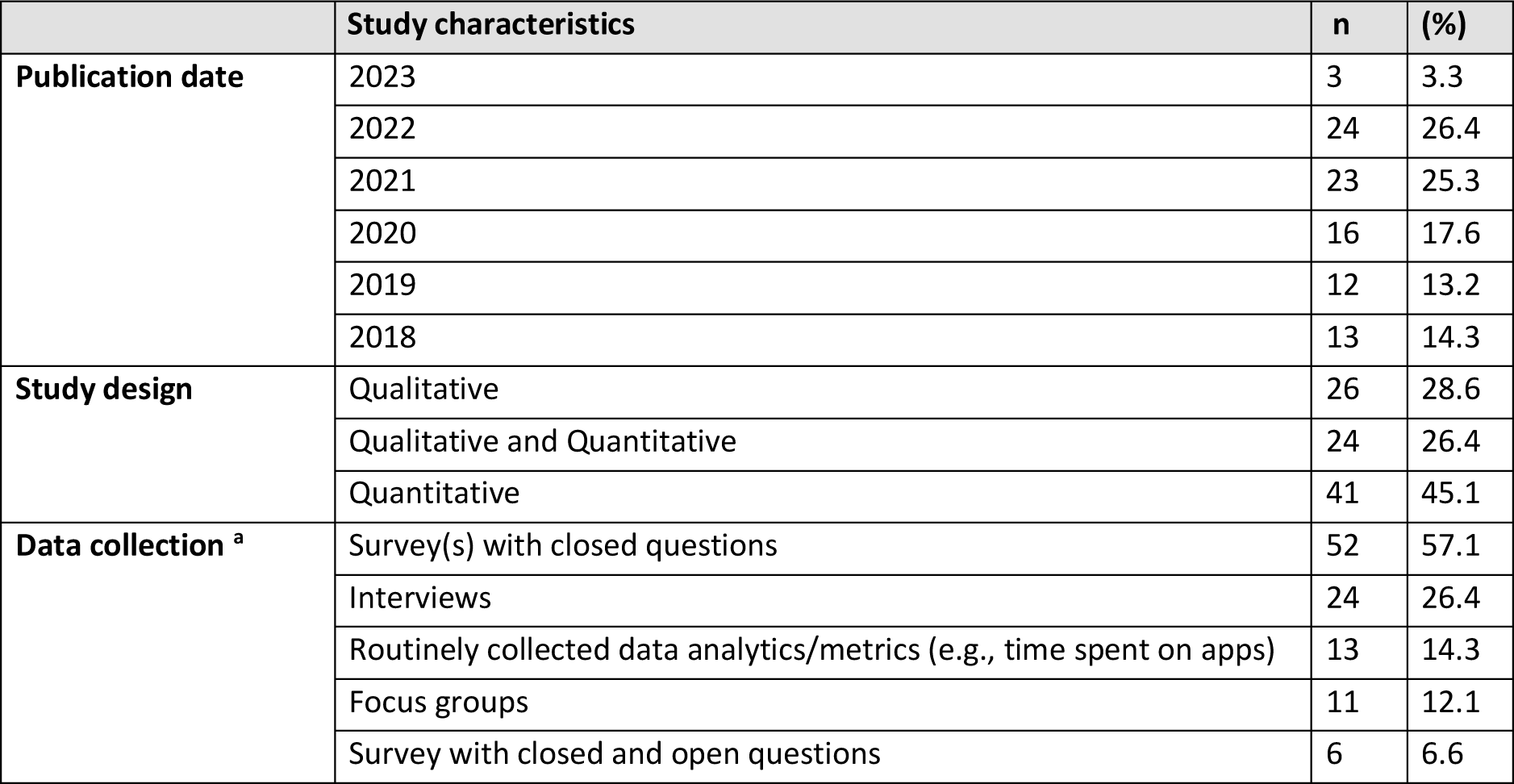
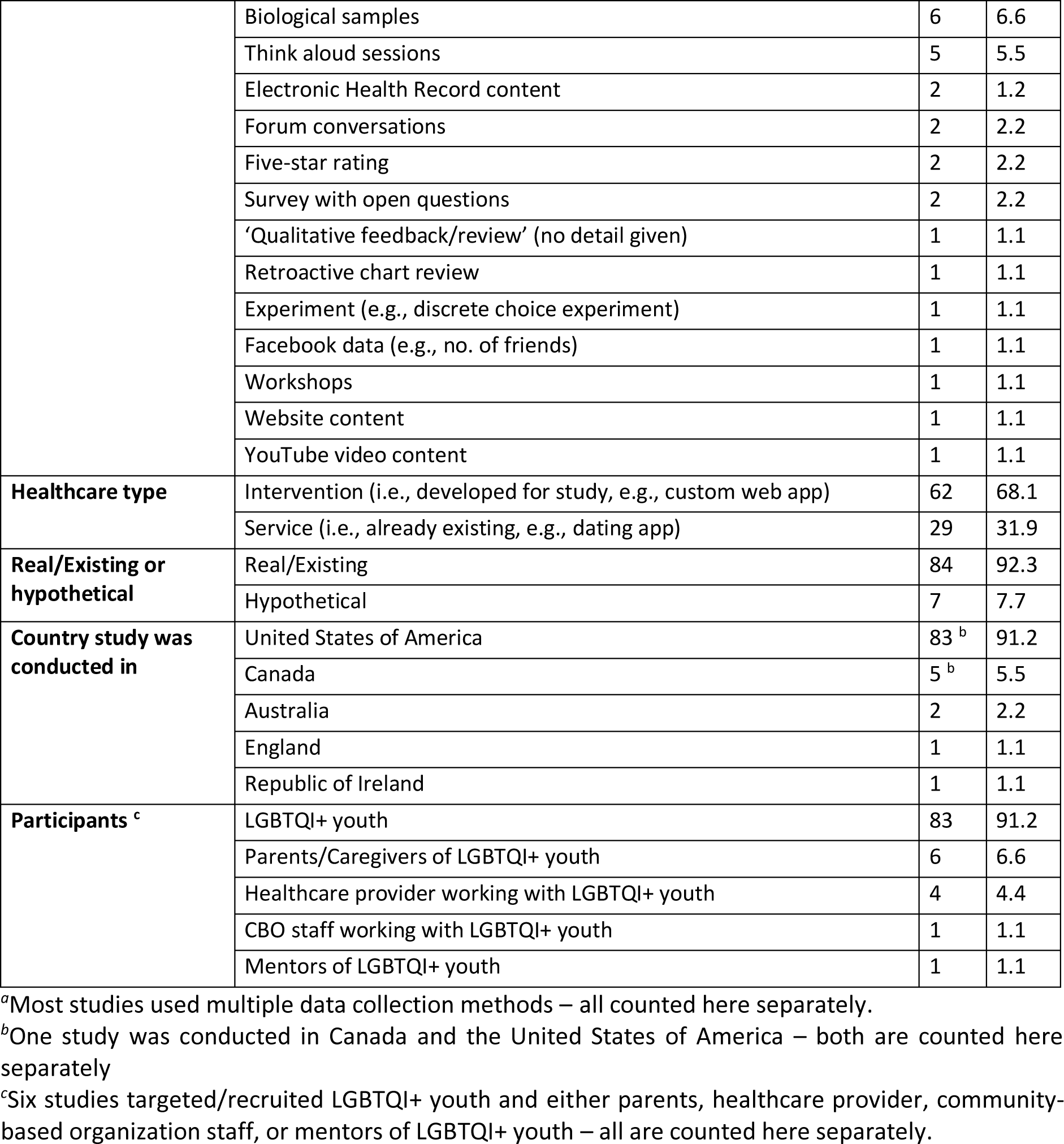
Study characteristics of included papers in the scoping review.

### RQ1: What types of online sexual and reproductive healthcare and transgender healthcare have received attention for LGBTQI+ youth and where are there gaps?

Table 3 provides an overview of which areas of heath and subsequent healthcare types have received attention regarding online healthcare for LGBTQI+ youth. In total, 76/91 papers explored sexual and reproductive health. The majority of the papers focused on sexual health (71/91, 78%), 3/91 explored reproductive health (3%), and 2/91 explored sexual and reproductive health together (2%). Further, one paper was categorised as exploring and sexual health and transgender health, as it examined use of the internet for both sexual health and transgender health information as well as online technologies for HIV management. However, it is important to note that this paper did not explore integrated SRHC and transgender healthcare. The remaining 15/91 focused on transgender health (17%).

**Table 3.** Types of online sexual and reproductive healthcare and transgender healthcare explored for LGBQTI+ youth in published literature.

| Area of health (n) | Health outcomes | n | Healthcare type |  |  |  |  |  |
| --- | --- | --- | --- | --- | --- | --- | --- | --- |
|  |  |  | Education/ Information e.g., | n | Non-clinical Support e.g., | n | Clinical care e.g., | n |
| **Sexual health (71 <sup>a</sup>)** | STI and BBV prevention<br>*STIs and BBV (n=21)*<br>*HIV only (n=25)*<br>*Human papillomavirus (HPV) only (n=6)* | 52 | HIV risk reduction; HIV testing; PrEP; How to have conversations about sex with healthcare provider and partners; HIV disclosure; STIs; STI risk reduction; STI testing; Condom use/ effectiveness; Coping with minority stress; Interpersonal and substance related risk factors; Safe sex; Sex pressures; HPV risk and prevalence; HPV vaccination and effectiveness; HPV vaccination cost and insurance | 42 | Reminders to get tested for HIV/ take PrEP; Q&A with healthcare provider; Personalized HIV risk reduction/ PrEP adherence strategies; PrEP adherence tracking; Feedback on PrEP adherence; Skill development for HIV disclosure; GPS maps for finding STI and HIV test sites; Reminders to get tested for STIs and HIV/ take PrEP; Skill building for condom use; Reminders to get vaccinated for HPV; GPS maps for finding HPV vaccination sites; Links to resources (LGBTQI+ friendly providers); Q&A with healthcare provider | 35 | Home delivery of HIV self-test kits; eConsultation with healthcare provider for PrEP; Home delivery of STI and HIV self-sample kits; Home delivery of condoms; eConsultation with healthcare provider for completing tests; Home delivery of PrEP | 10 |
|  | HIV management | 9 <sup>a</sup> | AIDS and HIV; Adherence, retention, and self-management; How often to see healthcare provider; ART (names, common side effects, doses per day) | 7 | Reminders to take ART medication; Feedback on ART adherence; Attention training; Q&A with trained professionals or automated response | 8 |  | 0 |
|  | Sexual health *per se* | 9 <sup>a</sup> | Sexual health Relationships; Sex; How to access inclusive HIV testing; How to have conversations about sex; Safe sex | 9 | Building skills for having conversations about sex with parents and healthcare provider; Peer communication/ support/ advice regarding sexual health and safe sex | 4 |  | 0 |
|  | HIV stigma reduction | 2 | HIV stigma reduction | 1 | Peer discussions for stigma reduction and community building; Q&A with healthcare provider | 2 |  | 0 |
| **Reproductive health (3)** | Pregnancy prevention | 3 | Sex education; Birth control | 2 | Links to resources; Q&A with trained professional; Peer communication/ support | 2 |  | 0 |
|  | Reproductive care for cancer survivors | 1 | Providing reproductive care for adolescent and young adult LGBTQ cancer survivors | 1 |  | 0 |  | 0 |
| **Sexual and reproductive health (2)** | Sexual health and reproductive health *per se* | 1 | Sexual health; Sex education; Birth control, IUD, implant, the pill; Human Papilloma Virus prevention | 2 |  | 0 |  | 0 |
| **Transgender health (16 <sup>a</sup>)** | Gender identity/transition *per se* | 8 | Trans people's experiences; Gender affirming language | 7 | Peer support and communication; Coping skills for stigma; Links to resources | 5 |  | 0 |
|  | Gender affirming care (8) | 8 | Gender affirming care | 1 |  | 0 | eConsultation with healthcare provider for hormone replacement therapy | 8 |
| **Total** |  |  |  | 72 |  | 56 |  | 18 |

<sup>a</sup>Numbers within 'Area of health' equal 92 instead of 91 as one paper was categorized as sexual health and transgender health separately. Further, Numbers within 'Health outcomes' equal 93 instead of 91 as the same paper was subsequently categorized as both HIV management and sexual health *per se* separately within sexual health and gender identity/transition *per se* within transgender health.
<sup>b</sup>The majority of studies explored information/education, support, and/or clinical care, thus studies have been counted multiple times cross these three columns.

Regarding healthcare types, most of the papers explored provision of or engagement with education/information (72/91) and non-clinical support (56/91). Types of support included peer communication (e.g., forum discussions or social media interactions) (22/56); skill building (e.g., how to use condoms or have conversations about sex) (17/56); medication reminders (e.g., reminders to take PrEP) (13/56); Question and answer (Q&A) with a trained professional or with automated responses (11/56); service locators (e.g., for STI/HIV testing or PrEP) (11/56); personalised recommendations (e.g., tailored HIV testing frequency and PrEP use or strategies for ART adherence) (6/56); counselling (e.g., couples counselling for HIV testing and risk reduction) (3/56); and mentorship for HIV support (1/56) (135). Only 18/91 explored clinical care, most frequently telehealth/medicine (e-consultations with healthcare provider) (12/18) for gender affirming care (8/12) and STI/HIV prevention (4/12), home delivery of STI/HIV self-sample/self-test kits (9/18), home delivery of condoms (3/18), and (hypothetical) home delivery of PrEP (1/18). Table 2 depicts the details of online sexual, reproductive, and transgender healthcare for LGBTQI+ youth.

There were 10 different types of online platforms used across the 91 papers, including 9 papers in which it was unclear (i.e., web-based or eHealth, with no further description) and 1 paper categorized as ‘digital technologies’. The most frequently used online platforms were mobile apps (21/91, 23%), websites/web apps (17/91, 19%), telehealth/telemedicine which referred to video calls, typically with healthcare provider (12/91. 13%), SMS text (9/91, 10%), and social media (7/91, 8%). Figure 2 shows the online platforms used.

**Figure 2.**
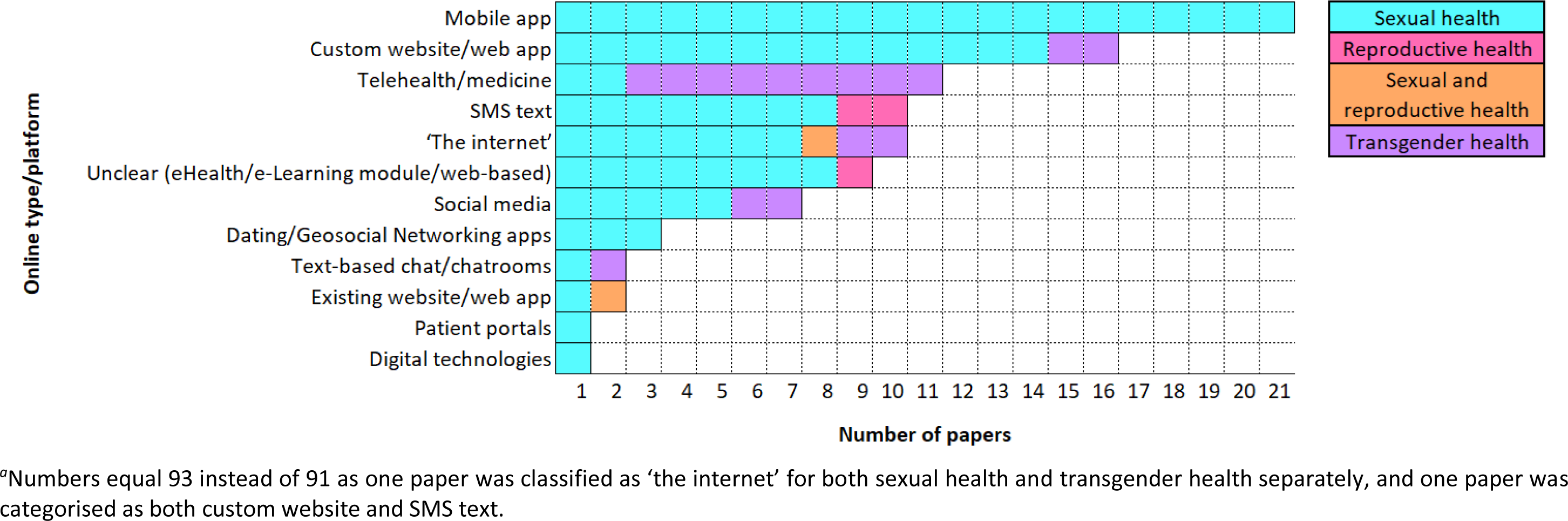
Online platforms used in sexual, reproductive, and transgender healthcare for LGBQTI+ youth.

Key gaps were research into health outcomes other than STI and BBV prevention, including sexual wellbeing and sexual violence/abuse; clinical care, including partner notification and management; reproductive health, including fertility preservation and assistance; education/information for transgender health, including gender affirming care; and integrated/combined SRHC and transgender healthcare.

### RQ2: Who are the target LGBTQI+ youth populations of online sexual and reproductive healthcare and transgender healthcare research, what additional intersectional factors have been considered, and where are there gaps?

The most frequently targeted LGBTQI+ population were GBMSM (62/91, 68%) (exclusively for sexual health), of which 26/62 were specific to cisgender men and 19/62 specified ‘assigned male at birth’ but were inclusive of people who identified as a different gender (e.g., non-binary) - 30/62 did not specify sex assigned at birth or gender identity (i.e., “gay men”). 27/91 papers targeted trans and gender diverse youth, almost half of which were for transgender health (14/27) or sexual health (HIV prevention) for trans women (9/28). 6/91 papers broadly targeted lesbian, gay, and bisexual (LGB or sexual minority) youth (1/6) or LGBT+ (sexual and gender minority) youth (5/6). Very few papers targeted sexual minority women (4/91), of which 2/4 were specific to cisgender women for reproductive health (pregnancy prevention); 1/4 specified gender inclusive ‘assigned female at birth’ and 1/4 was inclusive of people who identify as a woman, both of which were for sexual health. Tables 3 shows the LGBTQI+ youth populations targeted in online SRHC and transgender healthcare research by areas of health and health outcomes.

Across the 91 papers, targeted LGBTQI+ youth ranged in age from 7-36 years with 39 age ranges, the most common of which were 18-30 (9/91), 14-18 (7/91), 18-25 (6/91), 13-19 (5/91), 16-24 (5/91) and 18-24 (5/91) years. Almost half (41%) of youth targeted were aged 18 or older. Eight papers did not report a target age range nor age range of the participants. Figure 3 depicts target age ranges of LGBTQI+ youth targeted for online SRHC and transgender healthcare research.

**Figure 3.**
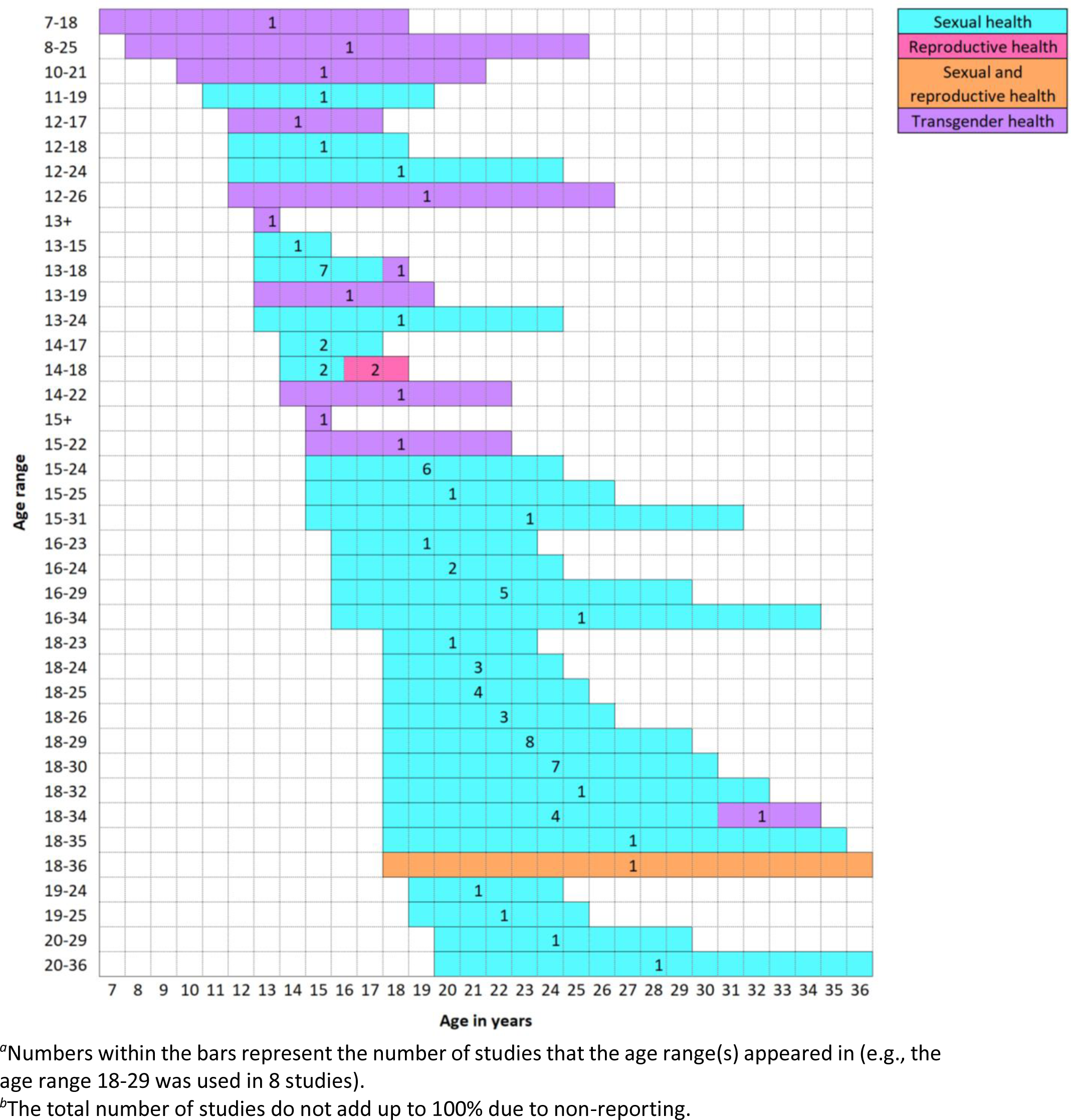
Target age ranges of LGBTQI+ youth for online sexual and reproductive healthcare and transgender healthcare.

Regarding intersectionality, in line with the remit of the current scoping review (i.e., ‘LGBTQI+ youth’), the most commonly identified PROGRESS+ variables were ‘Age’, considered in all of the papers, followed by ‘Gender/Sex’ (89/91), and ‘Sexual orientation/identity/behaviour’ (73/91). Other PROGRESS+ variables were Race/Ethnicity (25/91) for sexual healthcare (22/25), reproductive healthcare (1/25), sexual and reproductive health (1/25), and transgender healthcare (1/25); Place of Residence (5/91), Features of Relationships (5/91), Disability (1/91), and Social Networks (1/91), all for sexual health; Education (2/91) for reproductive health; and Living with HIV (10/91) largely for sexual health (HIV management). No papers considered Religion, Occupation, Socio-economic Status (i.e., income), or Time-dependent relationships in inclusion criteria for recruitment. Table 4 shows PROGRESS+ criterion considered in recruitment of target LGBTQI+ youth for online SRHC and transgender healthcare by healthcare dimensions and health outcomes.

**Table 4.** Target LGBQTI+ youth populations for online sexual and reproductive healthcare and transgender healthcare.

| Area of health (n) | Health outcome (n) | Target lesbian, gay, bisexual, queer/questioning, intersex, and other sexual and gender minorities (LGBTQI+) population (n) |  |  |  |  |  |  |  |  |  |  |  |
| --- | --- | --- | --- | --- | --- | --- | --- | --- | --- | --- | --- | --- | --- |
|  |  | LGBTQI+/<br>Sexual<br>and<br>gender<br>minority | Lesbian,<br>Gay,<br>Bisexual/<br>Sexual<br>minority | GBMSM<sup>b</sup><br>(sex/<br>gender not<br>specified) | GBMSM<br>(cisgender) | GBMSM<br>(gender<br>inclusive<br>assigned<br>male at birth) | GBMSM and<br>trans women<br>who have sex<br>with men | Trans and<br>gender<br>diverse | Trans<br>masc | Trans<br>women | Sexual<br>minority<br>women<br>(cis) | Sexual minority<br>assigned female<br>at birth<br>(gender inclusive<br>e.g., non-binary) | Sexual<br>minority<br>women<br>(sex<br>inclusive) |
| **Sexual health**<br>**(71)<sup>a</sup>** | STI and BBV prevention (21) | - | - | 6 | 9 | 5 | - | - | 1 | - | - | - | - |
|  | HIV only prevention (25) | - | - | 8 | 5 | 6 | 4 | 1 | - | 1 | - | - | - |
|  | HPV only prevention (n=6) | - | - | 4 | 2 | - | - | - | - | - | - | - | - |
|  | HIV management (9)<sup>a</sup> | - | - | 4 | - | 1 | 3 | - | - | 1<sup>a</sup> | - | - | - |
|  | SH *per se* (n=9)<sup>a</sup> | 1 | 1 | 1 | 1 | 1 | - | 2 | - | 1<sup>a</sup> | - | - | 1 |
|  | HIV stigma reduction (2) | - | - | - | - | 2 | - | - | - | - | - | - | - |
| **Reproductive health (3)** | Pregnancy prevention (2) | - | - | - | - | - | - | - | - | - | 2 | - | - |
|  | Reproductive care for cancer survivors (1) | 1 | - | - | - | - | - | - | - | - | - | - | - |
| **Sexual and reproductive health (2)** | Sexual and reproductive health *per se* (2) | 1 | - | - | - | - | - | - | - | - | - | 1 | - |
| **Transgender health (16)<sup>a</sup>** | Gender identity/transition *per se* (8)<sup>a</sup> | 2 | - | - | - | - | - | 5 | - | 1<sup>a</sup> | - | - | - |
|  | Gender affirming care (8) | - | - | - | - | - | - | 8 | - | - | - | - | - |
| **Total** |  | **5** | **1** | **23** | **17** | **15** | **7** | **17** | **1** | **2<sup>a</sup>** | **2** | **1** | **1** |

<sup>a</sup>Numbers within the cells equal 93 instead of 91 as one paper (115) (targeting trans women) was classified as both sexual health and trans health separately, and subsequently, HIV management and sexual health *per se* within sexual health and gender identity/transition *per se* within trans health. Thus, one paper targeting trans women is represented three times, however, this is not reflected in the total number of trans women, where the paper is only counted once.
<sup>b</sup>Gay, bisexual, and other men who have sex with men

Key gaps were research into online SRHC for populations other than GBMSM, in particular young sexual minority women and trans and gender diverse youth; and consideration of demographics and characteristics associated with inequalities in health, such as Place of Residence, Occupation, Religion, Education, and Socio-Economic Status.

**Table 5.** Intersectional factors considered in target LGBTQI+ youth populations for online sexual and reproductive healthcare and transgender healthcare.

| Area of health (n) | Health outcome (n) | PROGRESS+ variables considered when targeting LGBTQI+ youth for online sexual and reproductive healthcare and transgender healthcare |  |  |  |  |  |  |  |  |  |  |  |  |
| --- | --- | --- | --- | --- | --- | --- | --- | --- | --- | --- | --- | --- | --- | --- |
|  |  | Place of residence | Race/ Ethnicity | Occupation | Gender/ Sex | Religion | Education | Socio-economic status (income) | Social network | Features of relationships | Age | Sexual orientation/ identity/ behaviour | Disability | Living with HIV |
| **Sexual health (71) <sup>a</sup>** | STI and BBV prevention (21) | 3 | 5 | - | 21 | - | - | - | - | 3 | 21 | 21 | - | - |
|  | HIV only prevention (25) | 1 | 11 | - | 23 | - | - | - | 1 | 2 | 25 | 24 | - | - |
|  | HPV only prevention (6) | - | - | - | 6 | - | - | - | - | - | 6 | 6 | - | - |
|  | HIV management (9) <sup>a</sup> | 1 | 3 | - | 9 | - | - | - | - | - | 9 <sup>a</sup> | 8 | 1 | 9 <sup>a</sup> |
|  | SH *per se* (9) <sup>a</sup> | - | 1 | - | 8 <sup>a</sup> | - | - | - | - | - | 9 <sup>a</sup> | 6 | - | 1 <sup>a</sup> |
|  | HIV stigma reduction (2) | - | 2 | - | 2 | - | - | - | - | - | 2 | 2 | - | - |
| **Reproductive health (3)** | Pregnancy prevention (2) | - | 1 | - | 2 | - | 2 | - | - | - | 2 | 2 | - | - |
|  | Reproductive care for cancer survivors (1) | - | - | - | 1 | - | - | - | - | - | 1 | 1 | - | - |
| **Sexual and reproductive health (2)** | Sexual and reproductive health *per se* (2) | - | 1 | - | 2 | - | - | - | - | - | 2 | 2 | - | - |
| **Transgender health (16) <sup>a</sup>** | Gender identity/ transition *per se* (7) | - | 1 | - | 8 <sup>a</sup> | - | - | - | - | - | 8 <sup>a</sup> | 2 | - | 1 <sup>a</sup> |
|  | Gender affirming care (8) | - | - | - | 8 | - | - | - | - | - | 8 | - | - | - |
| **Total** |  | **5** | **25** | **0** | **89** | **0** | **2** | **0** | **1** | **5** | **91 <sup>a</sup>** | **74** | **1** | **10** |

<sup>a</sup>One paper was categorized as both sexual health and trans health separately, and subsequently HIV management and sexual health *per se*. Thus, one paper represented three times within the Gender/Sex column and the Age column. This is not reflected in the total numbers for these columns, where the paper is only counted once.

### RQ3: How, if at all, have theories, models, and frameworks have been used in research into online sexual and reproductive healthcare and transgender healthcare for LGBTQI+ youth?

Of the 91 included papers, 54/91 reported use of at least one framework, across which 44 frameworks were identified (see Table 6 for an overview by areas of health). Most papers reported only one framework 39/54; 15/54 papers reported more than one framework (two frameworks, 10/16; three frameworks, 3/16; four frameworks, 1/16; five frameworks, 1/16). Two of the 54 papers reported having used a framework without specifying which it was. A small number of papers reported using frameworks to provide a contextual lens for understanding how social structures influence people’s experiences (7/54) (e.g., Critical Race Theory or Intersectionality; (129)). Most of the studies reported using a framework(s) in an applied manner, for example, for development of study materials or intervention content, or to guide analyses or evaluation (50/54). However, 20/50 of these reported their study, intervention, or analysis to be ‘theory-driven’ (e.g., 76,113,120) or ‘informed by’ (e.g., (131,141), or having used a framework(s) to ‘guide the project’ or ‘for an in-depth exploration’ with no clear replicable explanation of how this was done – not including studies that specified this detail was published elsewhere. A key gap here is clear reporting of how theories are used.

**Table 6.** The theories, models, and frameworks, henceforth framework(s), used in online sexual and reproductive healthcare and transgender healthcare for LGBTQI+ youth.

| Health dimension (n) | Grouping (n) | Theory/Framework/Model | n | How Theory/Framework/Model have been used in studies (n) |  |  |
| --- | --- | --- | --- | --- | --- | --- |
|  |  |  |  | Theoretical | Applied | Unclear |
| **Sexual health (43)** | Communication (1) | Narrative Communication/Storytelling | 1 | - | 1 | - |
|  | Education/ Learning (2) | Dual Coding Theory | 1 | - | 1 | 1 |
|  |  | Entertainment Education | 1 | - | 1 | - |
|  |  | Cognitive Theory of Multimedia Learning | 1 | - | 1 | 1 |
|  | Healthcare (2) | Chronic Care Model | 1 | - | 1 | 1 |
|  |  | Patient-Centered Medical Home Model | 1 | - | 1 | 1 |
|  | Implementation (2) | Intervention Mapping | 1 | - | 1 | - |
|  |  | The RE-AIM Model | 1 | - | 1 | 1 |
|  | Mental Health (3) | Minority Stress Model | 1 | - | 1 | 1 |
|  |  | Resilience Framework | 1 | - | 1 | - |
|  |  | Stigma Theory | 1 | - | 1 | - |
|  | Social/Cognition (39) | Contingency Management | 1 | - | 1 | - |
|  |  | Dual Process Theory | 1 | - | 1 | - |
|  |  | Erdelez Model of Information Encountering | 1 | - | 1 | - |
|  |  | Information-Motivation-Behaviour Skills Model | 11 | - | 11 | 3 |
|  |  | Integrated Behavioural Model | 1 | - | 1 | - |
|  |  | Kari Conceptualization of Information Use | 1 | - | 1 | - |
|  |  | Social-Cognitive Theory | 9 | - | 9 | 2 |
|  |  | Social Norms Theory | 1 | - | 1 | - |
|  |  | Social-Personal (Theoretical) Framework | 2 | - | 2 | 2 |
|  |  | The Fogg Behavioural Model of Persuasive Technology | 1 | - | 1 | - |
|  |  | The Implementation Intention Theory | 1 | - | 1 | - |
|  |  | The Protection Motivation Theory | 3 | - | 3 | 3 |
|  |  | The Theory of Planned Behaviour | 3 | - | 3 | - |
|  |  | The Theory of Normative Social Behaviour | 1 | - | 1 | - |
|  |  | The Transtheoretical Model | 1 | - | 1 | - |
|  |  | The Wilson Model of Information Behaviour | 1 | - | 1 | - |
|  | Social Identity (1) | Social Identity Theory | 1 | - | 1 | - |
|  | Social/Structural (6) | Empowerment (Education) Theory | 2 | - | 2 | 1 |
|  |  | Gidden's Structuration Theory | 1 | 1 | 1 | - |
|  |  | Intersectionality | 1 | - | 1 | 1 |
|  |  | The Pharmacopornographisation Framework | 1 | 1 | - | - |
|  |  | The Social Ecological Model | 1 | - | 1 | 1 |
|  | Technology (3) | Information System Success Model | 1 | - | 1 | - |
|  |  | The Health Information Technology Usability Evaluation Model | 1 | - | 1 | - |
|  |  | The Unified Theory of Acceptance and Use of Technology Model | 1 | - | 1 | - |
|  | Unknown (2) | Not reported | 2 | - | 2 | 2 |
| **Reproductive health (3)** | Social/Cognition (1) | Information-Motivation-Behaviour Skills Model | 2 | - | 2 | - |
|  | Social/Structural (2) | Critical Race Theory | 1 | 1 | - | - |
|  |  | Intersectionality | 1 | 1 | - | - |
| **Transgender health (7)** | Education/ Learning (2) | Gagne's '9 External Events of Instruction' | 1 | - | 1 | - |
|  |  | Constructive Alignment Model | 1 | - | 1 | - |
|  | Healthcare (1) | The Iowa Model of Evidence-Based Practice | 1 | - | 1 | 1 |
|  | Implementation (1) | The RE-AIM Model | 1 | - | 1 | 1 |
|  | Mental Health (5) | Interpersonal Theory of Suicide | 1 | 1 | 1 | 1 |
|  |  | Minority Stress Model | 3 | 3 | 2 | 1 |
|  |  | Transactional Model of Stress and Coping | 1 | 1 | - | 1 |
|  | Social/Structural (2) | Intersectionality | 1 | 1 | 1 | 1 |
|  |  | Transgender Studies | 1 | - | 1 | 1 |
| **Sexual and reproductive health (1)** | Social/Structural (1) | The Structural Influence Model of Health Communication | 1 | 1 | - | - |

<sup>a</sup>Numbers do not add up to 100% as many studies used more than one framework.

## Discussion

### Principal results

The objective of this scoping review was to identify and describe existing literature on online SRHC and transgender healthcare for LGBTQI+ youth, synthesize study findings, and make recommendations for future research. This paper is the first to map a high volume of studies within online SRHC and transgender healthcare for LGBTQI+. The key findings were that most of the research was for sexual health, particularly HIV and STI prevention and HIV management. The majority of this research centered around the provision of or engagement with information/education and non-clinical support (e.g., reminders to get tested for HIV) and targeted young GBMSM aged 16 or 18 years and over (up to 36 years). There was little research into clinical care for sexual health (e.g., home delivery of STI/HIV testing kits, condoms, or PrEP). Additionally, there was very little research into reproductive health, with only two papers focusing solely on pregnancy prevention, two on general sexual and reproductive health, and one on the delivery of inclusive reproductive care for cancer survivors. Further, for transgender healthcare, half of the research focused on information/education and non-clinical support (e.g., peer communication/support) and the other half centered around telehealth/medicine (i.e., eConsultations) with healthcare provider regarding gender affirming care. The vast majority of this research targeted trans and gender diverse youth between ages 7 and 26. Moreover, there were a wide range of online platforms explored in researched, most of which were developed for a novel intervention including mobile apps, websites/web apps, and some of which were existing services such as websites, social media, and GSN/dating apps. Also, most research did not consider socio-economic demographics associated with inequalities in health (e.g., race/ethnicity, place of residence, occupation, and education (22,23) in the targeting of digital healthcare for LGBTQI+ youth. Finally, most papers used at least one theory or framework, indicating that the majority of interventions developed for online SRHC for LGBQTI+ youth are theory based.

### Comparison of key gaps in the literature with prior work and recommendations for future research

This scoping review demonstrates that there are key gaps in the literature. First is the dearth of research into online SRHC and transgender healthcare for LGBTQI+ youth outside the USA, particularly UK and Ireland. This is an important gap in research as financial cost is an often-reported barrier to SRHC and gender affirming care in the USA (146), given that their healthcare system is significantly more expensive than other countries (147), whereas, this is not a key issue in the UK due to the National Health Service providing healthcare free at the point of use, funded by public taxes (148). Therefore, conclusions drawn from studies regarding acceptability and barriers to online SRHC and gender affirming care for LGBTQI+ from USA may not be entirely applicable to the UK. Future research is needed to fill this gap and conduct research regarding online SRHC and transgender healthcare for LGBTQI+ outside the USA.

Another gap is research into health outcomes other than STI and BBV prevention, including sexual wellbeing and sexual violence/abuse. While these may fall under sexual health (149–151), neither were specified in any of the sexual healthcare papers included in this study. Further, these aspects of sexual health may be accessed by LGBTQI+ on online platforms designed for the general public, rather than LGBTQI+ youth (e.g., 28,150). However, research has shown that LGBTQI+ youth can struggle to identify relevant information and legitimate sources of information online (28,152). Moreover, despite sexual health, particularly STI and BBV prevention being the most researched, there was no literature regarding online options for partner notification and management which is problematic, as this is vital for reducing the spread of STIs (153).

A further gap was research into reproductive healthcare. The little research into online reproductive care largely focused on pregnancy prevention exclusively targeting cisgender sexual minority women yet LGBTQI+ youth, including but not limited to sexual minority women, are among the highest at risk for early and unplanned pregnancy (1,154,155). Additionally, LGBTQI+ youth may require fertility preservation or assistance (2,156–158) yet there was no research into this. Future research should explore the delivery and engagement with online healthcare for more expansive reproductive and fertility issues for LGBTQI+ youth.

Additionally, a critical gap was the lack of studies exploring education/information regarding gender affirming care for trans youth. The one study that explored this provided no detail about what the education provided via virtual visits entailed (60). This is an important finding as the internet, particularly social media, is a common and popular source of information of transgender healthcare for trans and gender diverse youth (159) yet they are lacking in official resources about transgender healthcare that have been rigorously developed. This also means that there is a lack of empirical research into how LBTQI+ youth are engaging with information about gender affirming care. Future research should consider development of formal educational resources for LGBTQI+ about gender affirming care.

Another gap is research into integrated/combined SRHC and transgender healthcare. This is of particular importance as SRHC integrated with transgender healthcare can facilitate access to both (50,51) and online care can overcome key barriers to SRHC and gender affirming care, delivering increased accessibility, convenience, and privacy (60,86,132,136,139). Together, this indicates that the provision of integrated SRHC and transgender healthcare has the potential to increase uptake of SRHC and improve sexual and reproductive health outcomes among trans youth. Yet, no studies have explored this. Future research should investigate the acceptability and feasibility of integrated care for LGBTQI+ youth.

A further vital gap is research into online sexual healthcare for populations other than GBMSM. While GBMSM have a disproportionately high burden of STIs/BBV (6), TDG youth and bisexual girls/women are also at high risk (1) and are considerably under-researched for digital innovations within sexual health prevention and management. Moreover, we found that socio-economic demographics and characteristics associated with inequalities in health (e.g., Place of Residence, Race/Ethnicity, Occupation, Religion, Education, Socio-Economic Status) (22,23) are largely overlooked when developing targeted digital SRHC interventions for LGBQTI+ youth. This is problematic as such demographics can impact access to online technology and the internet (160) and risk of poorer sexual and reproductive health outcomes (161–163). Future research should consider generating target sampling frames using the PROGRESS+ framework for purposeful recruitment of diverse populations.

Finally, there is an important gap regarding the reporting of the use of theory. As theory-based interventions support understanding of behaviour and behaviour change and have a higher likelihood of success, it is important that interventions are both theory and evidence based (164). While almost half of the papers in this scoping review reported applying a theory to the development of interventions (see Table 6), many of these did not report how this was done; they reported that the intervention was ‘theory-based’, for example, with no further explanation of what this entailed. Many papers did report use of theory to a high and replicable standard (e.g., 68,117,121). Future research could use these papers as an example of reporting the use of theory.

### Strengths

First, the volume of included papers in this scoping review provided a comprehensive overview of the literature into SRHC and transgender healthcare, depicting the breadth of research and identifying clear gaps (165). Further, following the JBI methodology (54) and using nine databases (166) for the search ensured that this scoping review was conducted in a rigorous and systematic manner and facilitated a thorough identification and mapping of the literature into online SRHC and transgender healthcare for LGBTQI+ youth, represented by the volume of studies. Another strength of the study was the contribution of two reviewers to screening both titles/abstracts and full texts. This reduced the chance of bias (167) and ensured that the eligibility criteria were well understood, and methods replicable, by a researcher outside the field of sexual health.

### Limitations

The volume of papers in this study classified it as a large scoping review and limited the detail that could be explored and cross paper analyses that could be conducted (165), such as thematic analysis to identify barriers and facilitators, due to the variety in participants and concepts (see S2 Appendix). Another limitation of this study may be that only ten papers’ titles, abstracts, and key words were used to specify search terms. However, no guidance on how many papers to include for identifying search terms is provided by JBI (53). Additionally, due to the volume of included papers, the reference lists of included papers were not searched. Therefore, while the authors’ best efforts were made to ensure that all possibly relevant studies were included in this scoping review, some papers may have been missed.

### Conclusions

While there is a wide range of research into online SRHC and trans healthcare, the majority of the existing research for SRHC focusses on the perspectives of young GBMSM pertaining to HIV and STI prevention and centers around the provision or engagement with education or information and non-clinical support, such as reminders to get tested or to take anti-retroviral medication. There are critical gaps in the literature including research focusing on reproductive healthcare, the provision of clinical care, and the perspectives of other LGBTQI+ sub-populations such as trans and gender diverse youth and young sexual minority women and women who have sex with women. Further, intersectional demographics and characteristics associated with inequalities in health such as education, occupation, income, religion, are chronically under-considered in recruitment. The PROGRESS+ framework could be a useful tool for targeted recruitment of diverse populations. Given that there is a shift to the delivery of healthcare online and LGBTQI+ youth have disproportionately poor sexual health outcomes and low engagement with SRHC, it is vital that these research gaps are filled to ensure that LGBTQI+ are considered and included in the development and delivery of online healthcare.

## Abbreviations, Acronyms, and Initials

GBMSM: Gay, bisexual, and other men who have sex with men
HIV: Human immunodeficiency virus
HPV: Human papillomavirus
LGBTQI+: Lesbian, gay, bisexual, transgender, queer, questioning, intersex, and other sexual orientation and gender diverse and minority populations
PCC: Participants, Concept, and Context
PrEP: Pre-exposure prophylaxis
SRHC: Sexual and reproductive healthcare
STI: Sexually transmitted infection
Trans: Transgender and gender diverse
UK: United Kingdom
UN: United Nations
USA: United States of America
WHO: World Health Organisation

## Data Availability

The full dataset for the present review is available online on Open Science Framework (OSF)

https://osf.io/ktwxn

## Acknowledgements

We would like to acknowledge the contribution of Ron O’Kane (RO), a PhD student at GCU, for acting as the second reviewer. We would also like to acknowledge Ruth Leiser, a Research Associate at the University of Strathclyde, for peer reviewing this scoping review.

## Funding

This scoping review was funded by a GCU PhD scholarship awarded to the first author (JMcL), who is a member of the Sexual Health and Blood Borne Viruses (SHBBV) research group and the Research Centre for Health (ReaCH). Supervisors for the PhD, JMacD and CSE, are supported by GCU. PF, based at the University of Strathclyde, and JG, based at University College London, are supported by their host institutions, as supervisors of the PhD.

## S1 Appendix

**S1 Appendix:** Glossary of terms used within this paper, their definitions, and references.

| Term | Definition |
| --- | --- |
| Cisgender | Those whose gender identity aligns with their sex assigned at birth (168,169). |
| Gender | The socially constructed, not biologically determined, norms and roles associated with masculinity and femininity (e.g., wearing specific clothes) (39). |
| Gender affirmation | A social/appearance and/or physical/medical transition to align one's gender expression and body with their gender identity (39,170). |
| Gender affirming care | Clinical care to treat gender dysphoria and/or support people in living in the gender that is most authentic and comfortable to them (e.g., mental health assessments; puberty blockers; menstrual suppression; hormone replacement therapy; fertility preservation or assistance, gender reassignment surgery) (48,49,171–175). |
| Gender diverse and minority populations | Those whose gender identify and expression do not conform to societal norms associated with their birth-assigned sex including but not limited to transgender, intersex, non-binary, gender neutral, gender fluid, gender non-conforming (168,169). |
| Gender dysphoria | Emotional distress due to incongruence between sex and gender identity (40). |
| Gender expression | The way in which people manifest, convey and present their gender (170). |
| Gender identity | Internal sense of gender (170). |
| Heteronormative | The concept that heterosexuality is the preferred or normal mode of sexual orientation, assuming the gender binary (i.e., that there are only two distinct, opposite genders) (39). |
| Intersex | Those who may have the biological attributes of both sexes or whose biological attributes do not fit with societal assumptions about what constitutes male or female (169). |
| Medical transition | Taking hormones (e.g., testosterone or oestrogen) and/or undergoing surgery to align one's body with their gender identity (40,49,176,177). |
| Partner notification | When sex partners are informed of their exposure to an STI or HIV (178). |
| Pre-exposure prophylaxis (PrEP) | An antiretroviral drug to reduce the risk of acquiring HIV (179). |
| Queer | A term used by those wanting to reject specific labels of romantic orientation, sexual orientation and/or gender identity (168,169). |
| Questioning | Exploring one's sexual orientation and/or gender identity (168,169). |
| Sex | Biological characteristics, such as chromosomes and reproductive organs, traditionally associated with male and female; however, sex is not binary, given the significant overlap in sex characteristics between 'male' and 'female' people (39). |
| Sexual and reproductive health | An umbrella term for issues including the absence of illness or disease, such as STIs and HIV; pregnancy, fertility and reproductive matters; sexual safety, freedom from coercion, violence or abuse; and sexual wellbeing and the ability to express one's sexuality (151,180,181). |
| Sexual and reproductive health care (SRHC) | Information, advice and support, and clinical care for: 1) STI and HIV prevention, testing, and treatment; 2) fertility and pregnancy prevention, preservation, assistance, and termination; 3) sexual safety, abuse, and violence; and 4) sexual wellbeing (e.g., STI/HIV |
|  | testing; PrEP; partner notification; human papillomavirus vaccination; contraception; in vitro fertilization; intrauterine insemination; medication to prevent pregnancy; counselling) (182–189). |
| Sexual orientation diverse and minority populations | Those whose attraction to others is non-heterosexual including, but not limited to, lesbian, gay, bisexual, pansexual, queer, questioning (168,169). |
| Sexual wellbeing | Satisfaction with sexual relationships and functioning; comfort with sexuality; resilience and forgiveness regarding sexual experience; and sexual awareness, self-esteem, security, and respect (149,151). |
| Social transition | Altering one's gender expression, appearance, and other social aspects to match their gender identity, such as clothes, hair, name, pronouns (40,170). |
| Telemedicine | Video conferencing for conversations or counselling with a healthcare provider (123). |
| Trans | An umbrella term for gender diverse and minority populations (173). |
| Transgender health | Living in the gender that feels most authentic and comfortable, based on the premise that sex and gender are distinct. This can involve gender affirmation and transition to align one's body and gender expression with their gender identity (39). |
| Transgender healthcare | Information, support, and clinical care regarding gender identity, expression, and transition. Includes but is broader than gender affirming care (47) |
| Youth | Approximately ages 10-35 years; encompasses 'youth', 'young people', 'teens', 'adolescents', and 'young adults' (e.g., 1,28,63,190–194). |

## S2 Appendix

### S2 Appendix: Deviations from the protocol

There were four deviations from the protocol, listed below.

1. A quality appraisal was not conducted, as the high-volume of papers and heterogeneity of services led to an alteration of research questions regarding acceptability and barriers/facilitators to simply state the number of studies that explored these concepts, rather than conducted further analyses to make interpretations or draw conclusions about acceptability.
2. “and evaluate the methodological quality of” was removed from the objective, as a quality appraisal was no longer needed.
3. Four or the six original research questions (RQs) were altered and one was deleted (see S2 Table for an overview and rationale for each) – RQ6 remained unchanged but was renumbered as RQ3. All RQs were updated to refer to ‘transgender healthcare’ in place of ‘gender healthcare’ for a more accurate representation of the concept. Additionally, ‘developed’ was removed from all RQs – while this was still included in the eligibility criteria, this terminology is outdated.
4. A grey literature search was not conducted due to the unexpectedly high number of included studies from the database search – a grey literature search was deemed unnecessary given the high volume of peer-reviewed, published papers.

**S2 Table.** Deviations in research questions from protocol.

| Research question | Protocol | Paper | Rationale |
| --- | --- | --- | --- |
| RQ1 | What dimensions of digital SRHC and GHC for LGBTQI+ youth in high-income, developed countries have received attention in the literature and who are the target populations? | What types of online sexual and reproductive healthcare and transgender healthcare have received attention for LGBTQI+ youth and where are there gaps? | ‘and who were the target populations [of online sexual, reproductive and transgender healthcare]’ was removed, as this was better addressed in a separate research question (RQ2) in the results. |
| RQ2 | What are the characteristics of LGBTQI+ youth in research regarding digital SRHC and GHC and accessing and using digital SRHC and GHC in high-income, developed countries? | Who are the target LGBTQI+ youth populations of online sexual and reproductive healthcare and transgender healthcare research, which additional intersectional characteristics have been considered, and where are there gaps? | The question about who was ‘accessing and using’ digital SRHC and transgender healthcare was removed, as the data available did not support answering this. Additionally, the question about the ‘characteristics of LGBTQI+ youth in research’ was replaced by ‘who are the target LGBTQI+ youth populations in research’ and ‘what intersectional characteristics have been considered in recruitment’, as these questions better represent the author’s intentions to identify which LGBTQI+ youth populations were the target of digital healthcare services and interventions. |
| RQ3 | What is the acceptability of digital SRHC and GHC in high-income, developed countries for LGBTQI+ youth? | Deleted | Given the volume of final included papers, studies exploring acceptability, barriers, and facilitators were too heterogenous with regards to participants and concepts to conduct a cross-study analysis. Continuing with this analysis would have resulted in potentially misleading findings, given the diversity of LGBTQI+ populations, skewed focus on GBMSM, and variation in digital platforms and healthcare types. A follow up, more focused, systematic review would be more appropriate to address the original question. Detail on how many papers explored acceptability, barriers, and facilitators can be found in Appendix 7. This will be addressed in the final PhD thesis. |
| RQ4 | What are the barriers and facilitators to LGBTQI+ youth accessing and using digital SRHC and GHC in high-income, developed countries? | Deleted |  |
| RQ5 | How is LGBTQI+ ‘youth’ defined in digital SRHC and GHC research from high-income, developed countries? | Deleted | This question was anticipated to lack in research impact and age ranges were addressed under the research question about target populations. This will be addressed in the final PhD thesis. |
| RQ6 | How, if at all, have theory or frameworks been used in research into digital SRHC and GHC for LGBTQI+ youth in high-income, developed countries? | How, if at all, have theories, models, and frameworks been used in research into online sexual and reproductive healthcare and transgender healthcare for LGBTQI+ youth? |  |

## S3 Appendix

### S3 Appendix: Inclusion and exclusion criteria for eligibility by participant, concept, and context (PCC) and their rationale

Regarding Participants, LGBTQI+ included gender and sexual orientation diverse and minority identifying participants as the target population, inclusive of all terms to describe sexual minorities and gender diverse populations (e.g., non-binary, gender non-conforming, gender fluid, and gender neutral (168,169)). ‘Youth’ refers to the target populations of ‘youth’, ‘young people’, ‘young adults’ and ‘adolescents/teens’ and/or included participants within an age range of 10-35 years. This broad age range was selected to capture the maximum number of studies based on an initial search of online SRHC and transgender healthcare for LGBTQI+ youth research (e.g., 10-20, (191); 13-29, (1); 16-24, (63); 16-29, (28); 18-26, (192); 15-34, (194)).

Within Concept were four key components: online; sexual and reproductive health; transgender health; and healthcare. Here, ‘online’ encompassed all SRHC and transgender healthcare delivered via internet based digital technology including, but not limited to websites; web apps; mobile apps; short messaging service (SMS); email; and video calls (e.g., 195). Sexual and reproductive health included all aspects of sexual and reproductive health including infection and disease (e.g., STIs and BBVs), fertility and pregnancy, sexual wellbeing, and sexual safety, violence, and abuse (151,180,181). Transgender health included all aspects of gender identity, expression, transition, and affirmation (39). Moreover, taking an inclusive approach to ‘healthcare’, this included the delivery or engagement with any interventions or services aimed at preventing, treating, or managing illness or disease or promoting wellbeing related to sexual and reproductive health and transgender health for LGBTQI+ youth. Services refer to existing online SRHC or transgender healthcare for help-seeking, such as information on websites; advice or support via text-based conversations with peers or trained professionals such as bi-directional email, SMS text live chat (synchronous text-based chat platform), forums, social media groups; or clinical care, such as access to online STI/HIV self-sampling kits, partner notification, PrEP, contraception, counselling, or e-consultations (virtual/remote platforms for video or audio consultations with healthcare provider) (e.g., 11,16,19,46,196–198). Interventions refer to online strategies that have been developed to change a specific sexual and reproductive health or transgender health related behaviour(s) or outcome(s) for LGBTQI+ youth populations including education programmes to increase knowledge (e.g., 58,70,79,93,94,104,107,114,199), support for accessing or using services such as signposting to local services (e.g., 200), or novel provision of a digital version of a service that is typically delivered in person (e.g., 201–203).

Finally, for Context, this scoping review seeks to form a foundational understanding of how to optimize UK-based online SRHC for LGBTQI+ youth. As online SRHC and transgender healthcare and barriers to care can differ considerably between countries, depending on infrastructure and social welfare/protections for access to health care (204,205), we focus on countries with similar contexts to the UK. Therefore, studies from high-income and developed economy countries, as defined by the United Nations (UN) (57), were included.

See S3 Table for further detail and rationales.

**S3 Table.**
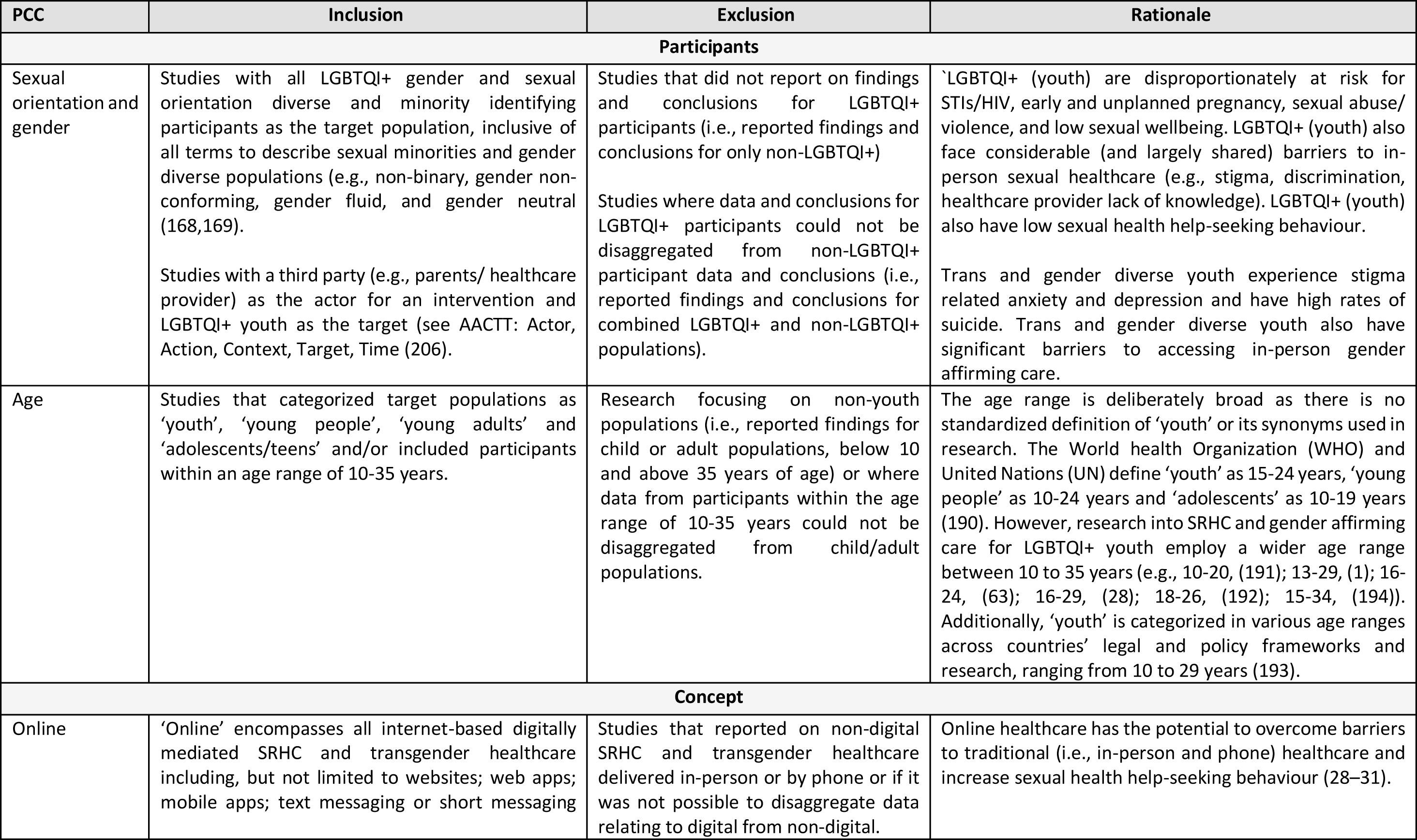
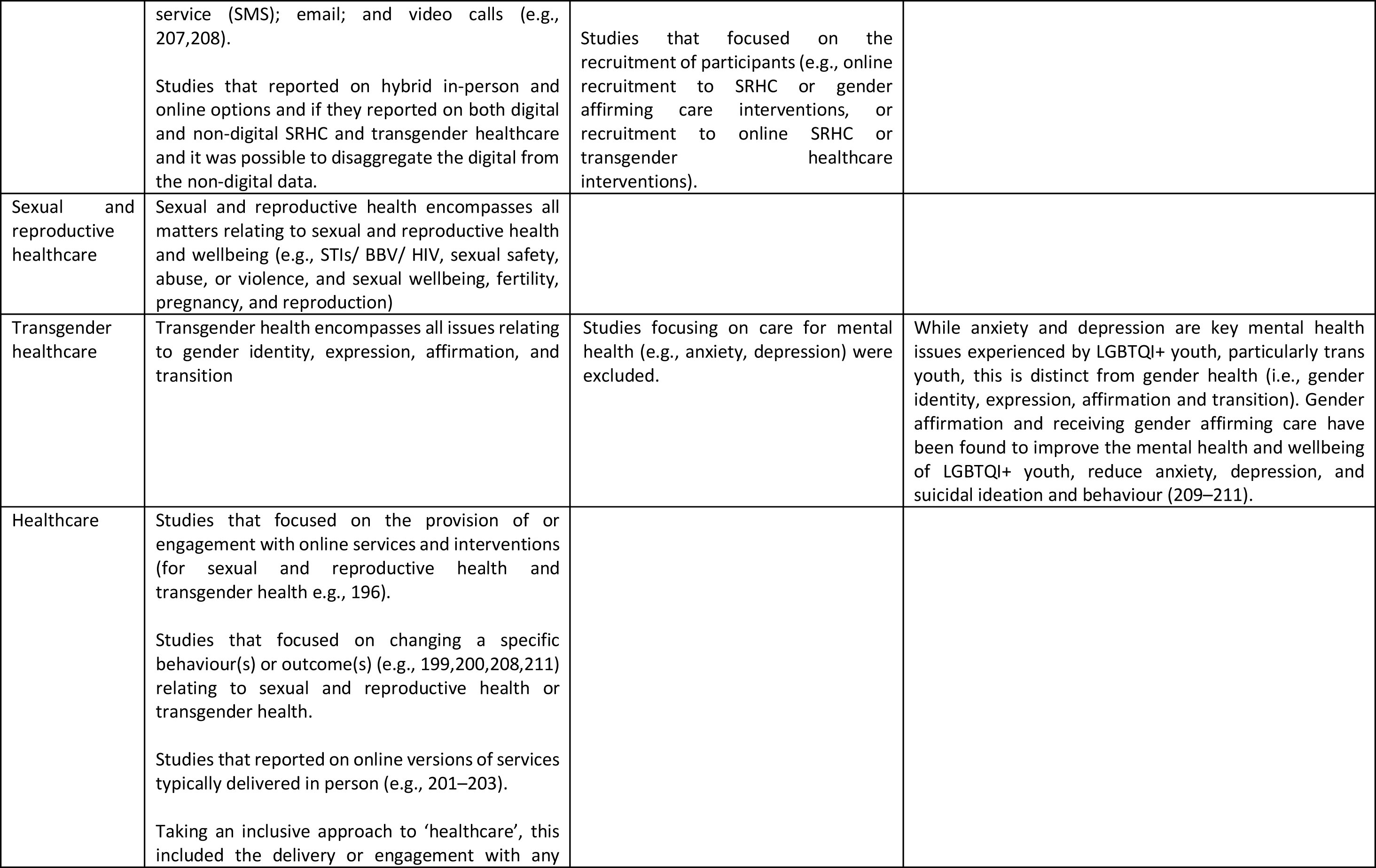
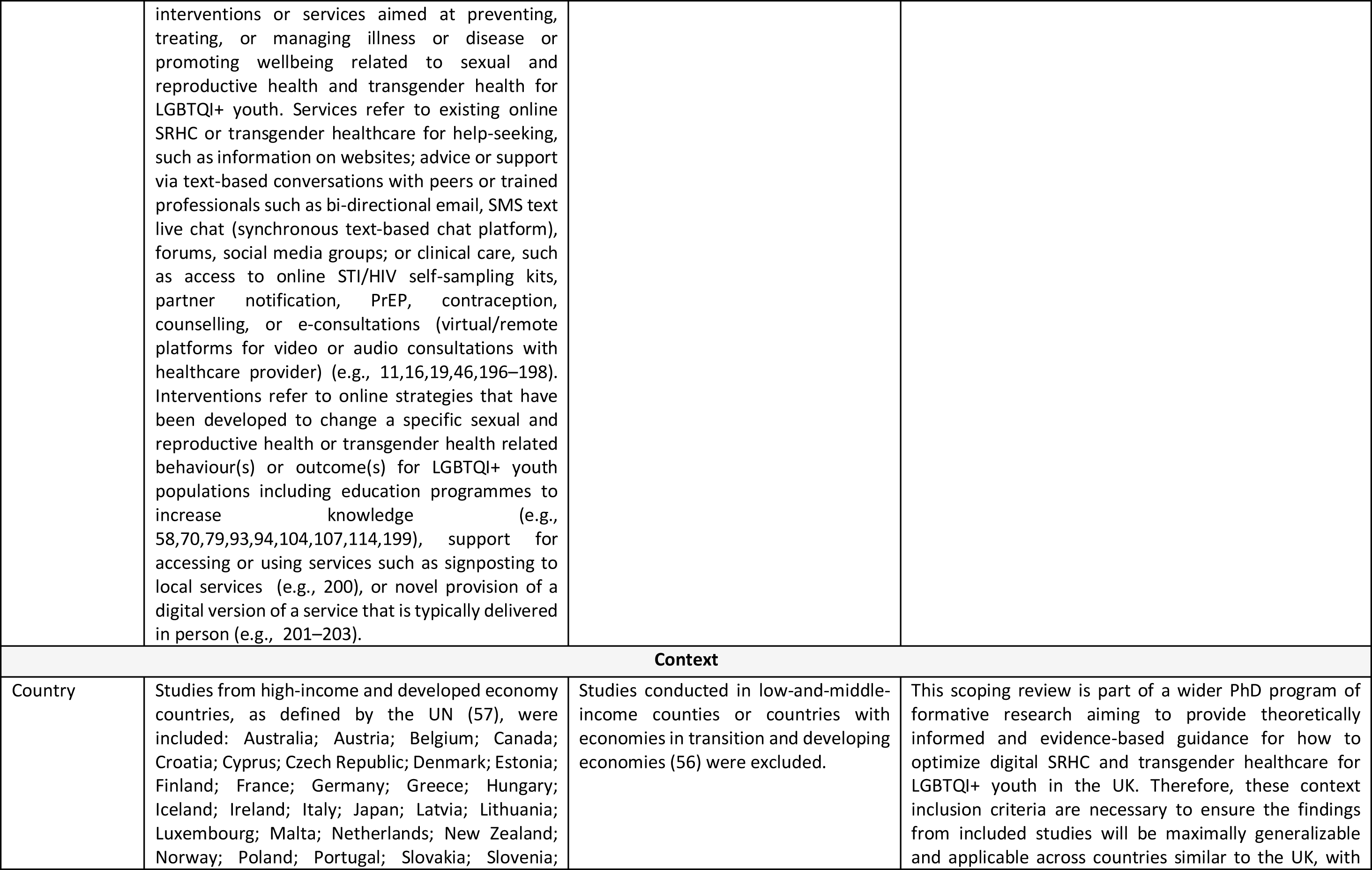
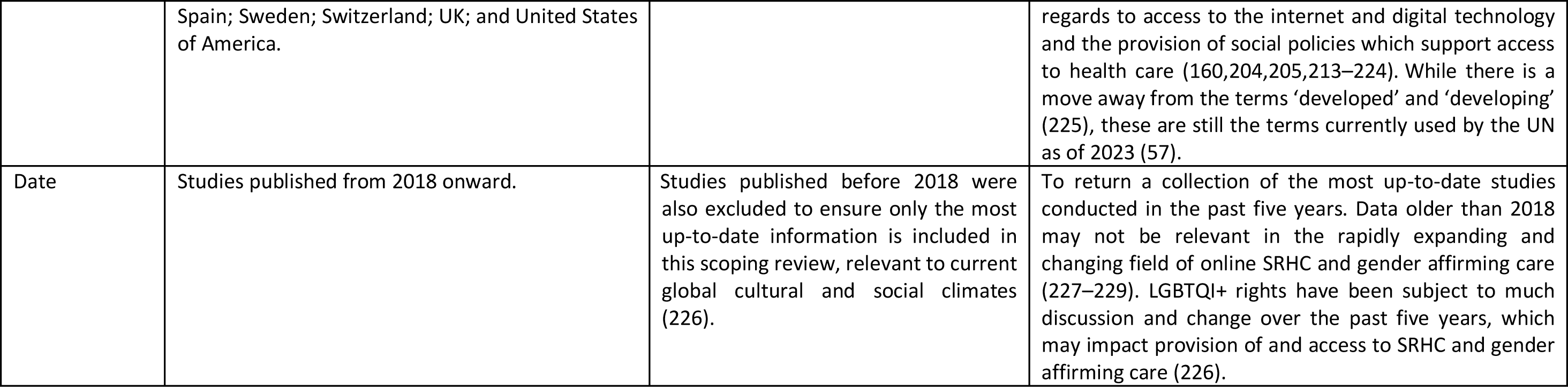
Inclusion and exclusion criteria for eligibility by participant, concept, and context (PCC) and their rationale.

| PCC | Inclusion | Exclusion | Rationale |
| --- | --- | --- | --- |
| **Participants** |  |  |  |
| Sexual orientation and gender | Studies with all LGBTQI+ gender and sexual orientation diverse and minority identifying participants as the target population, inclusive of all terms to describe sexual minorities and gender diverse populations (e.g., non-binary, gender non-conforming, gender fluid, and gender neutral (168,169).<br>Studies with a third party (e.g., parents/ healthcare provider) as the actor for an intervention and LGBTQI+ youth as the target (see AACCT: Actor, Action, Context, Target, Time (206). | Studies that did not report on findings and conclusions for LGBTQI+ participants (i.e., reported findings and conclusions for only non-LGBTQI+)<br>Studies where data and conclusions for LGBTQI+ participants could not be disaggregated from non-LGBTQI+ participant data and conclusions (i.e., reported findings and conclusions for combined LGBTQI+ and non-LGBTQI+ populations). | 'LGBTQI+ (youth) are disproportionately at risk for STIs/HIV, early and unplanned pregnancy, sexual abuse/ violence, and low sexual wellbeing. LGBTQI+ (youth) also face considerable (and largely shared) barriers to in-person sexual healthcare (e.g., stigma, discrimination, healthcare provider lack of knowledge). LGBTQI+ (youth) also have low sexual health help-seeking behaviour.<br>Trans and gender diverse youth experience stigma related anxiety and depression and have high rates of suicide. Trans and gender diverse youth also have significant barriers to accessing in-person gender affirming care. |
| Age | Studies that categorized target populations as 'youth', 'young people', 'young adults' and 'adolescents/teens' and/or included participants within an age range of 10-35 years. | Research focusing on non-youth populations (i.e., reported findings for child or adult populations, below 10 and above 35 years of age) or where data from participants within the age range of 10-35 years could not be disaggregated from child/adult populations. | The age range is deliberately broad as there is no standardized definition of 'youth' or its synonyms used in research. The World health Organization (WHO) and United Nations (UN) define 'youth' as 15-24 years, 'young people' as 10-24 years and 'adolescents' as 10-19 years (190). However, research into SRHC and gender affirming care for LGBTQI+ youth employ a wider age range between 10 to 35 years (e.g., 10-20, (191); 13-29, (1); 16-24, (63); 16-29, (28); 18-26, (192); 15-34, (194)). Additionally, 'youth' is categorized in various age ranges across countries' legal and policy frameworks and research, ranging from 10 to 29 years (193). |
| **Concept** |  |  |  |
| Online | 'Online' encompasses all internet-based digitally mediated SRHC and transgender healthcare including, but not limited to websites; web apps; mobile apps; text messaging or short messaging | Studies that reported on non-digital SRHC and transgender healthcare delivered in-person or by phone or if it was not possible to disaggregate data relating to digital from non-digital. | Online healthcare has the potential to overcome barriers to traditional (i.e., in-person and phone) healthcare and increase sexual health help-seeking behaviour (28–31). |
|  | service (SMS); email; and video calls (e.g., 207,208).<br>Studies that reported on hybrid in-person and online options and if they reported on both digital and non-digital SRHC and transgender healthcare and it was possible to disaggregate the digital from the non-digital data. | Studies that focused on the recruitment of participants (e.g., online recruitment to SRHC or gender affirming care interventions, or recruitment to online SRHC or transgender healthcare interventions). |  |
| Sexual and reproductive healthcare | Sexual and reproductive health encompasses all matters relating to sexual and reproductive health and wellbeing (e.g., STIs/ BBV/ HIV, sexual safety, abuse, or violence, and sexual wellbeing, fertility, pregnancy, and reproduction) |  |  |
| Transgender healthcare | Transgender health encompasses all issues relating to gender identity, expression, affirmation, and transition | Studies focusing on care for mental health (e.g., anxiety, depression) were excluded. | While anxiety and depression are key mental health issues experienced by LGBTQI+ youth, particularly trans youth, this is distinct from gender health (i.e., gender identity, expression, affirmation and transition). Gender affirmation and receiving gender affirming care have been found to improve the mental health and wellbeing of LGBTQI+ youth, reduce anxiety, depression, and suicidal ideation and behaviour (209–211). |
| Healthcare | Studies that focused on the provision of or engagement with online services and interventions (for sexual and reproductive health and transgender health e.g., 196).<br>Studies that focused on changing a specific behaviour(s) or outcome(s) (e.g., 199,200,208,211) relating to sexual and reproductive health or transgender health.<br>Studies that reported on online versions of services typically delivered in person (e.g., 201–203).<br>Taking an inclusive approach to ‘healthcare’, this included the delivery or engagement with any<br>interventions or services aimed at preventing, treating, or managing illness or disease or promoting wellbeing related to sexual and reproductive health and transgender health for LGBTQI+ youth. Services refer to existing online SRHC or transgender healthcare for help-seeking, such as information on websites; advice or support via text-based conversations with peers or trained professionals such as bi-directional email, SMS text live chat (synchronous text-based chat platform), forums, social media groups; or clinical care, such as access to online STI/HIV self-sampling kits, partner notification, PrEP, contraception, counselling, or e-consultations (virtual/remote platforms for video or audio consultations with healthcare provider) (e.g., 11,16,19,46,196–198). Interventions refer to online strategies that have been developed to change a specific sexual and reproductive health or transgender health related behaviour(s) or outcome(s) for LGBTQI+ youth populations including education programmes to increase knowledge (e.g., 58,70,79,93,94,104,107,114,199), support for accessing or using services such as signposting to local services (e.g., 200), or novel provision of a digital version of a service that is typically delivered in person (e.g., 201–203). |  |  |
| **Context** |  |  |  |
| Country | Studies from high-income and developed economy countries, as defined by the UN (57), were included: Australia; Austria; Belgium; Canada; Croatia; Cyprus; Czech Republic; Denmark; Estonia; Finland; France; Germany; Greece; Hungary; Iceland; Ireland; Italy; Japan; Latvia; Lithuania; Luxembourg; Malta; Netherlands; New Zealand; Norway; Poland; Portugal; Slovakia; Slovenia; | Studies conducted in low-and-middle-income countries or countries with economies in transition and developing economies (56) were excluded. | This scoping review is part of a wider PhD program of formative research aiming to provide theoretically informed and evidence-based guidance for how to optimize digital SRHC and transgender healthcare for LGBTQI+ youth in the UK. Therefore, these context inclusion criteria are necessary to ensure the findings from included studies will be maximally generalizable and applicable across countries similar to the UK, with |
|  | Spain; Sweden; Switzerland; UK; and United States of America. |  | regards to access to the internet and digital technology and the provision of social policies which support access to health care (160,204,205,213–224). While there is a move away from the terms ‘developed’ and ‘developing’ (225), these are still the terms currently used by the UN as of 2023 (57). |
| Date | Studies published from 2018 onward. | Studies published before 2018 were also excluded to ensure only the most up-to-date information is included in this scoping review, relevant to current global cultural and social climates (226). | To return a collection of the most up-to-date studies conducted in the past five years. Data older than 2018 may not be relevant in the rapidly expanding and changing field of online SRHC and gender affirming care (227–229). LGBTQI+ rights have been subject to much discussion and change over the past five years, which may impact provision of and access to SRHC and gender affirming care (226). |

## S4 Appendix

**S4 Appendix:**
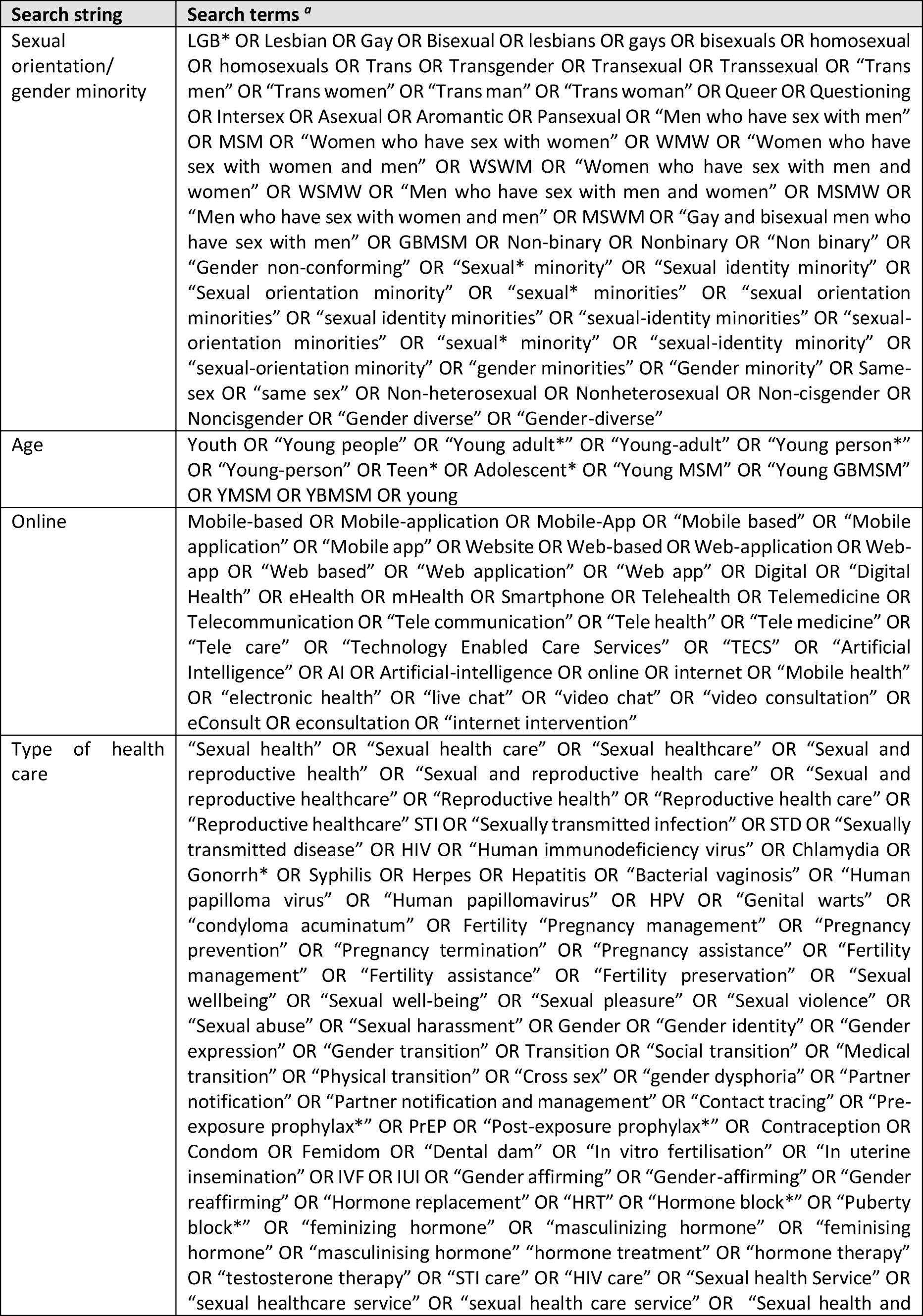
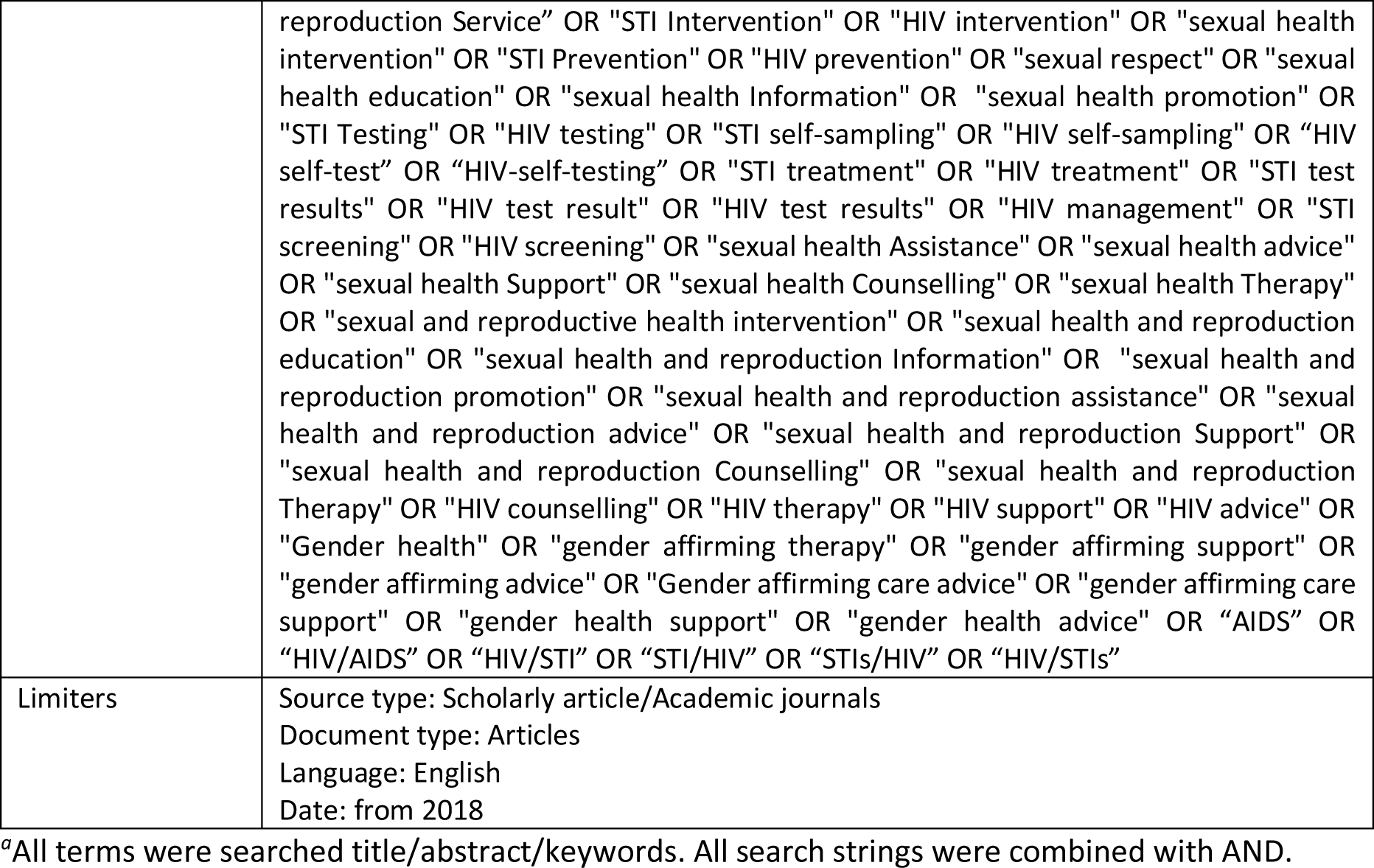
Search terms generated for MEDLINE (EBSCO).

## S5 Appendix

**S5 Appendix:** Data extraction tool adapted from JBI.

|  | Variables (columns in Excel) | Data extracted |
| --- | --- | --- |
| **Study details** | Author of publication | First author's last name, et al. |
|  | Date of publication |  |
|  | Study aim/objective | Verbatim from the abstract or introduction |
|  | Study research questions/ objectives | Verbatim from the introduction |
|  | Data collection methods | Verbatim or deductive from the methods (categories: Quantitative; Qualitative; Qualitative and Quantitative) |
|  | Data collection method details | Verbatim from the methods |
|  | Analysis methods | Verbatim from the methods |
|  | Sample size | Verbatim from the methods |
|  | Eligibility/Inclusion criteria | Verbatim from the methods |
|  | Recruitment methods | Verbatim from methods (including sampling type, where participants were recruited from, how they were recruited) |
| **RQ1 (Concept)** | Area of health | Deductive based on focus of the study from the title and aim/objective (categories: Sexual Health; Reproductive health; Sexual and Reproductive Health; Transgender health) |
|  | Health outcome | Inductive, based on the focus of the study, from the title, aim/objective and methods (categories: HIV prevention; STI prevention; HIV and STI prevention; HIV management; HIV stigma reduction; Pregnancy prevention; Sexual health (per se); Reproductive health (per se); Gender identity/expression; and Gender affirmation/transition) |
|  | Health care type: Education/ Information | Deductive from the title, aim/objective and methods (refers to services and interventions that impart information only typically to achieve an increase in knowledge (e.g., information about STIs/HIV; contraception; or gender expression and transition). |
|  | Health care type: Education/ Information type | Verbatim from the methods explaining the intervention/service |
|  | Health care type: Non-clinical Support | Deductive from the title, aim/objective and methods (refers to services and interventions that provide non-clinical emotional or practical support, beyond information, typically to achieve a desired outcome (e.g., peer communication for HIV stigma reduction; reminders for increased PrEP adherence; or skill building for increased skills for partner notification). |
|  | Health care type: Non-clinical support type | Verbatim from the methods explaining the intervention/service |
|  | Health care type: Clinical care | Deductive from the title, aim/objective and methods (refers to services and interventions for medical care, specific to testing, diagnosing, treating, and managing sexual and reproductive health issues or |
|  |  | gender affirming care (e.g., STI/HIV testing; uptake or maintenance of PrEP; consultations for gender affirming hormones). |
|  | Health care type: Clinical care type | Verbatim from the methods explaining the intervention/service |
|  | Online type | Verbatim from the title, abstract, aim, and/or methods explaining the intervention/service |
|  | Intervention/service | Deductive from the methods describing the intervention/service (categories: Intervention; Service) |
|  | Real/Hypothetical | Deductive from the methods describing the intervention/service (categories: Intervention; Service) |
|  | Intervention name | Verbatim from title, introduction, aim, or methods |
|  | Intervention/service details | Verbatim from the methods describing the intervention/service |
| **RQ2**<br>**(Participants)** | Target LGBTQI+ population | Verbatim from title, aim, and/or methods |
|  | Target LGBTQI+ population age range | Verbatim from introduction, aim, or methods |
|  | PROGRESS+: Place of residence | Deductive from recruitment or eligibility/inclusion criteria in the methods (categories: N; Y with verbatim details) |
|  | PROGRESS+: Race/ Ethnicity | Deductive from recruitment or eligibility/inclusion criteria in the methods (categories: N; Y with verbatim details) |
|  | PROGRESS+: Occupation | Deductive from recruitment or eligibility/inclusion criteria in the methods (categories: N; Y with verbatim details) |
|  | PROGRESS+: Gender/Sex | Deductive from recruitment or eligibility/inclusion criteria in the methods (categories: N; Y with verbatim details) |
|  | PROGRESS+: Religion | Deductive from recruitment or eligibility/inclusion criteria in the methods (categories: N; Y with verbatim details) |
|  | PROGRESS+: Education | Deductive from recruitment or eligibility/inclusion criteria in the methods (categories: N; Y with verbatim details) |
|  | PROGRESS+: Socio-economic status (income) | Deductive from recruitment or eligibility/inclusion criteria in the methods (categories: N; Y with verbatim details) |
|  | PROGRESS+: Social network | Deductive from recruitment or eligibility/inclusion criteria in the methods (categories: N; Y with verbatim details) |
|  | PROGRESS+: Age | Deductive from recruitment or eligibility/inclusion criteria in the methods (categories: N; Y with verbatim details) |
|  | PROGRESS+: Disability | Deductive from recruitment or eligibility/inclusion criteria in the methods (categories: N; Y with verbatim details) |
|  | PROGRESS+: Sexuality | Deductive from recruitment or eligibility/inclusion criteria in the methods (categories: N; Y with verbatim details) |
|  | PROGRESS+: Features of relationships | Deductive from recruitment or eligibility/inclusion criteria in the methods (categories: N; Y with verbatim details) |
|  | PROGRESS+: Time dependent relationships | Deductive from recruitment or eligibility/inclusion criteria in the methods (categories: N; Y with verbatim details) |
|  | Other: Living with HIV | Deductive from recruitment or eligibility/inclusion criteria in the methods (categories: N; Y with verbatim details) |
|  | Target descriptor category | Verbatim from title, aim, and/or methods (categories: youth; young [LGBTQI+]; young adults; young people; teen; adolescent – from) |
|  | Participant age range | Verbatim from methods |
|  | Participant age stats | Verbatim from methods (including mean and standard deviation; median and interquartile range; age range percentages) |
| **RQ3** | Theory/Model/Framework | Verbatim from introduction or methods |
|  | How Theory/Model/Framework was used | Verbatim and summarized from the introduction, methods, results, or discussion (refers to any/all detail on the Theory/Model/Framework) |
|  | Descriptive/Social lens | Inductive from the introduction, methods, results, or discussion (categories: Y; N) |
|  | Predictive/Applied | Inductive from the introduction, methods, or results (categories: Y; N) |
|  | Unclear | Inductive from the introduction, methods, or results (categories: Y; N) |
|  | Number of Theory/Model/Frameworks used | Calculated from description of Theory/Model/Frameworks in introduction or methods |
| **Context** | Country | Verbatim from methods or inductive from introduction or methods |

<sup>a</sup>Where a study did not report a data point, this was extracted as N/R (not reported).

For RQ1, data were extracted and analyzed regarding areas of health, health outcomes, types of healthcare, and online platforms explored. For areas of health, papers were deductively categorized as belonging to sexual, reproductive, or transgender health. For health outcomes, data were inductively identified from the aim or methods of the paper. For types of healthcare, data were deductively categorized as Education/Information, Non-clinical Support, and Clinical Care. Education/Information refers to services and interventions that impart information only typically to achieve an increase in knowledge (e.g., information about STIs/HIV; contraception; or gender expression and transition). Non-clinical Support refers to services and interventions that provide non-clinical emotional or practical support, beyond information, typically to achieve a desired outcome (e.g., peer communication for HIV stigma reduction; reminders for increased PrEP adherence; or skill building for increased skills for partner notification). Clinical care refers to services and interventions for medical care, specific to testing, diagnosing, treating, and managing sexual and reproductive health issues or gender affirming care (e.g., STI/HIV testing; uptake or maintenance of PrEP; consultations for gender affirming hormones). Finally, for online platforms, the verbatim terms used in papers by authors were extracted. For online platforms, data were extracted from the title, research aims, or methods using the verbatim terms used in papers by authors.

For RQ2, data were extracted and analyzed regarding which LGBTQI+ populations were targeted for the intervention/service and which intersectional factors were considered in eligibility or recruitment. For target LGBTQI+ populations, data were extracted from the title or methods using the verbatim terms used in papers by authors. For analysis, different terms with the same or similar meanings were combined, for example, ‘same sex attracted boys/men’, ‘gay men’, ‘men who have sex with men’, were categorized as GBMSM. Moreover, the PROGRESS+ framework (PROGRESS and other factors associated with inequalities in health outcomes, including Sexual orientation, Age, and Disability) (22,23) was used to identify how intersectionality was considered. We also included Living with HIV as an additional factor.

For RQ3, frequency counts and percentages were calculated for the number of papers that reported use of one or more theory, model or framework (hereby framework) and how they were used. In extraction, how frameworks were used was captured deductively, grouped into either ‘Theoretical’ or ‘Applied’. ‘Theoretical’ describes when frameworks were used to provide a contextual lens for understanding how social structures influence people’s experiences. ‘Applied’ describes when frameworks were used in an applied manner, for example, for development of study materials or intervention content, or to guide analyses or evaluation.

## Notes

### Competing Interest Statement

The authors have declared no competing interest.

### Clinical Protocols

https://www.medrxiv.org/content/10.1101/2023.08.25.23294615v1

